# A systematic review of ontology-based clinical decision support system rules: usage, management, and interoperability

**DOI:** 10.1101/2022.05.11.22274984

**Authors:** Xia Jing, Hua Min, Yang Gong, Dean F. Sittig, Paul Biondich, David Robinson, Tim Law, Adam Wright, Christian Nøhr, Arild Faxvaag, Lior Rennert, Nina Hubig, Ronald Gimbel

## Abstract

**Objective:** Clinical decision support systems (CDSS) have a critical role in improving the quality and safety of health care delivery. CDSS rules direct the behavior of CDSS. However, the CDSS rules have not been routinely shared and reused, and ontology can promote the reusing of CDSS rules. We systematically screened literature to elaborate on the current status of ontology applied in CDSS rule management.

**Methods:** We searched PubMed, the Association for Computing Machinery (ACM) Digital Library, and the Nursing & Allied Health Database for publications focusing on ontology, clinical decision support, and rules. Grounded theory and PRISMA 2020 guidelines were followed. One author started the screening and literature analysis, and two authors validated the processes and results. Inclusion and exclusion criteria were developed and refined iteratively.

**Results:** Among 81 included publications, the identified CDSS were mainly applied to managing chronic conditions, alerts for medication prescriptions, reminders for immunizations and preventive services, diagnoses, and treatment recommendations. The CDSS rules were presented in Semantic Web Rule Language, Jess, or Jena formats. Despite ontology was used to supply medical knowledge, CDSS rules, and terminologies to CDSS, ontology has not been used in CDSS rule management.

**Conclusions:** Although ontology can facilitate the reuse, management, and maintenance of CDSS rules, CDSS ontology remains unavailable indicating that more efforts are needed to improve the reusability and interoperability of CDSS rules.

## Introduction

Clinical Decision Support Systems (CDSS) have been studied and used in clinical care delivery for longer than half a century [1–5]. The effectiveness of CDSS in clinical care has been established [6–8]. We cite some prominent examples in CDSS. Middleton et al. reviewed the two decades of clinical decision support experiences as CDSS users and developers and discussed their vision of CDSS for the future [9]. Wright et al. recommended best practice guidelines in CDSS [10–14]. Meanwhile, Sittig and colleagues comprehensively documented the challenges of CDSS [15]. McDonald is one of the earliest pioneers who demonstrated the effectiveness of CDSS in clinical care [16]. Meeting clinicians’ information needs is one way to improve clinical care outcomes via CDSS. Many studies have shown the effectiveness of such CDSS, for example, Cimino’s Infobutton [6, 17]. Currently, CDSS is used routinely in clinical care in the USA. An analysis of a 2015 national survey showed CDSS usage rates reached 68.5% to 100% in primary care settings based in offices [18] in the USA. Therefore, CDSS is commonly used in clinical care delivery as part of electronic health record (EHR) systems. CDSS has a range of forms: such as reminders for preventive services (e.g., immunizations, screening tests) [19, 20], alerts for drug-drug interactions [14, 21, 22], diagnostic or treatment plan recommendations [23–25], content assistance for clinicians [26–30], or recommendations for adhering to current clinical practice guidelines [31–33]. CDSS has played a critical role and is used commonly in providing safer and better quality clinical care services.

CDSS rules, like central neural systems for humans, direct the behaviors of a CDSS during operations by incorporating a patient’s data, contextual information, and medical domain knowledge. CDSS rules have a central role and are a decisive factor in the relevance and usefulness of CDSS in the entire clinical workflow, including whether a CDSS will be adopted and used. CDSS rules can be written in different languages, such as Arden syntax [34], the Semantic Web Rule Language (SWRL), Jess, Jena, and other programming languages. Reusing and sharing CDSS rules is ideal since it is very costly to recreate these rules every time, but the process is not a routine operation across organizations. As a component of an EHR system, updating the CDSS rules regularly is needed to keep the CDSS relevant and useful in clinical care delivery. However, it is a very time-consuming and resource-demanding process to create, manage, maintain, reuse, and share CDSS rules—one type of CDSS artifact. This resource-demanding process has been recognized by larger institutes [4, 35], which usually are better resourced than small-scale practices. Therefore, creating and maintaining CDSS rules can be much more challenging in resource-limited office-based practices.

The ontology can facilitate the use, reuse, sharing, and interoperability of CDSS rules, solving the challenges of managing and maintaining CDSS rules across institutional borders. Ontology is the enabling technology of the Semantic Web [36] and is critical in information sharing and reusing [37, 38]. Although there are many definitions of ontology, we use the one by Gruber: “an ontology is a specification of conceptualization” [37]. Ontology has been used to generate and supply domain knowledge in many fields, including medicine [39] and CDSS [40], which is not an ontology that focuses on CDSS but includes relevant content of CDSS. Ontology can be used to shed light on achieving interoperable clinical information, such as CDSS rules, at a more granular level.

Although ontology can facilitate the reuse and sharing of CDSS rules, no CDSS ontology exists. Therefore, we are building a CDSS ontology [41]. Furthermore, to elaborate on the current status of using ontology in CDSS rules, to improve CDSS rules **reuse and interoperability**, we systematically reviewed existing English literature about the **intersection of CDSS rules**, Semantic Web technology (especially **ontology)**, and the use of ontologies in CDSS. The primary **rationale** of this systematic literature review is that even though there is no existing CDSS ontology except for our effort, the systematic examination of the literature on the intersection of CDSS rules, ontology, and the use of ontology in CDSS can provide a clearer picture on how ontology is used in CDSS, especially regarding CDSS rules. The literature review can shed light on our effort to use the Semantic Web technology to enable the reuse and maintenance of CDSS rules.

Through this effort, we aimed to achieve **three objectives**. First, this systematic review will provide an initial structure and considerations for organizing our CDSS ontology. Second, the results can help us hone our focus area: ontology use in CDSS rule management. A clear scope and comprehensive picture can form a knowledge framework of the topic that may inspire and guide future efforts to advance this field more explicitly. Third, the manually annotated results of selected publications will be used as gold standards to automatically identify entities from the literature, which complements efforts to curate CDSS ontology manually.

## Methods

### General workflow

After the literature search, the first 100 of all the retrieved papers were initially screened by one author (XJ), and initial inclusion and exclusion criteria were drafted. This screening was replicated, and the results were discussed, validated, and the criteria were refined and adjusted by two other authors (HM and YG). Therefore, the first 100 papers were screened by all three authors independently. The inclusion and exclusion criteria were developed, revised, and refined during the iterative literature screening, review, analysis, and discussions. Then the rest of the papers were screened by at least two authors (XJ and MH, or XJ and YG) independently to determine if a paper should be included in the literature review. The discrepancies were discussed and resolved via iterative rounds of teleconferences.

When needed, the literature was first screened using publication titles, abstracts, and full-text publications. Then after determining which papers to include, included papers were manually coded to provide more content analysis and synthesized evidence. The final results were shared and agreed upon among all authors. Therefore, the screening of each paper and manual review of included papers were conducted by two authors independently and approved by at least two authors first and all authors later. All discrepancies and concerns were resolved via group email discussions and complementary teleconferences. Figure 1 illustrates the general workflow.

**Figure 1.**
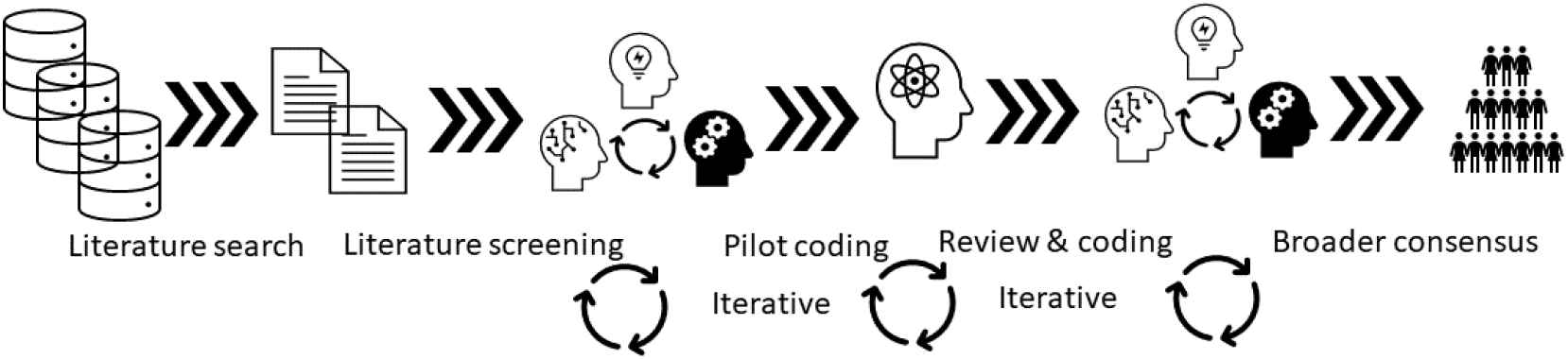
General workflow of the systematic literature review

### Databases and search strategies

On June 2, 2020, we did an initial set of literature searches. After initial review and discussions, we refined and agreed with the search strategies. On Sept 15, 2020, we searched PubMed, the Association for Computing Machinery (ACM) Digital Library, and the Nursing & Allied Health Database for literature with the following search strategies. On January 5, 2022, we conducted the exact search again in the three literature databases as an update.

- PubMed

(clinical decision support systems[MeSH Terms]) AND (ontolog*[Title/abstract] OR rule*[Title/abstract])

- ACM Digital Library

The following search was conducted within the scope of the ACM Guide to Computing Literature.

[[Publication Title: “clinical decision support*”] OR [Publication Title: cds*]] AND [[Publication Title: ontolog*] OR [Abstract: ontolog*] OR [Publication Title: rule*] OR [Abstract: rule*]]

- Nursing & Allied Health Database (NAHD)

The following search was limited to peer-reviewed publications.

mesh(clinical decision support) AND (ti(ontology) OR ti(ontologies) OR ab(ontology) OR ab(ontologies) OR ti(rule) OR ti(rules) OR ab(rule) OR ab(rules))

### Inclusion and exclusion criteria

The inclusion criteria were:

1. Literature is written in English;
2. Full-text publication available;
3. Ontology designed to implement or implemented in CDSS, especially related to CDSS rules;
4. Content includes granularity of CDSS rules;
5. Ontology designed to integrate or already integrated with health information systems (e.g., EHR), either in a production system or a prototype, with at least one architecture diagram;
6. Applied in clinical domains or designed for clinical domains to support health care providers;
7. Peer-reviewed publication; and
8. Contained details on integration between CDSS and EHR for evaluation studies.

The exclusion criteria were:

1. Only CDSS rules, regardless of any stage of the CDSS rules’ lifecycle (i.e., development, identification, refinement, validation, evaluation, or implementation), or no integration was mentioned, or no mention of ontology;
2. Only Ontology was developed, evaluated, and validated, or no integration was mentioned, or no CDSS was mentioned;
3. System was designed without mentioning the granularity of CDSS rules or ontology; and
4. The publication described nonclinical decisions, such as administrative or management decisions (e.g., supply chain management).

### Reviewing, coding, analyzing, and synthesizing processes

During the reviewing and manual coding, we followed grounded theory. One author (XJ) randomly selected ten papers from the *included* 81 papers to start the coding based on the focus of this literature review: CDSS rules (including application and management), ontology, and roles of ontology. The coding results were discussed by three authors (XJ, HM, and YG). The discussion results formed the first draft of codes and code groups (Appendix 1), i.e., data items. Then the three coders (authors) reviewed and coded the first 40 of the *included* papers using the initial principles and code groups and adding new codes and code groups when needed. Midway through coding, consensus on updated principles and code groups was obtained at a second meeting and following iterative discussions. Then the refined codes and code groups were used to code the remaining papers. Every paper was coded by at least two coders independently. Then the coding results were compared, and any discrepancies were discussed to resolve. During each discussion, the code groups and codes were revised, consolidated, and updated. The refined code groups and examples are described in Appendix 2. Therefore, the data items were emerged during review and refined via discussions instead of pre-defined before reviewing.

A qualitative data analytic tool-ATLAS.ti 9 (desktop version and Web version), was used for coding by the three coders. To form the final codes and code groups, all projects (ATLAS.ti organizes the data as projects, i.e., each coder has a project to save all the papers and coding results) from all three coders were merged, and any duplicates were removed after a manual check. Then codes and code groups were later combined, and any duplicates were removed after a manual review.

After coding, the literature was analyzed and synthesized. For this process, we focused on several aspects, including the application domains of CDSS, the mechanism of CDSS used in clinical settings, CDSS rule format, authoring, management, and ontology roles. The process was iterated several times among the three coders first. Then after obtaining consensus among the three coders, the results were shared and discussed among all authors later. Through iterative discussions, any concerns, confusion, or disagreements among authors were resolved with group emails and complementary teleconferences.

We have followed the PRISMA 2020 checklist [42] in reporting the systematic review with all relevant items.

## Results

### Results related to CDSS characteristics

The literature searches generated 1,235 publications from three sources until Jan 5, 2022. After removing duplicates and examining according to inclusion and exclusion criteria, we included 81 publications (Appendix 3) in the final review and analysis [19-21, 23-25, 43-117]. Figure 2 illustrates each step of the literature search, screening, selection flow, and results. Figure 3 summarizes the main components covered by the literature review and the summary findings. Figure 3 also serves as an initial knowledge framework on CDSS, CDSS rules, and ontology applications in CDSS.

**Figure 2.**
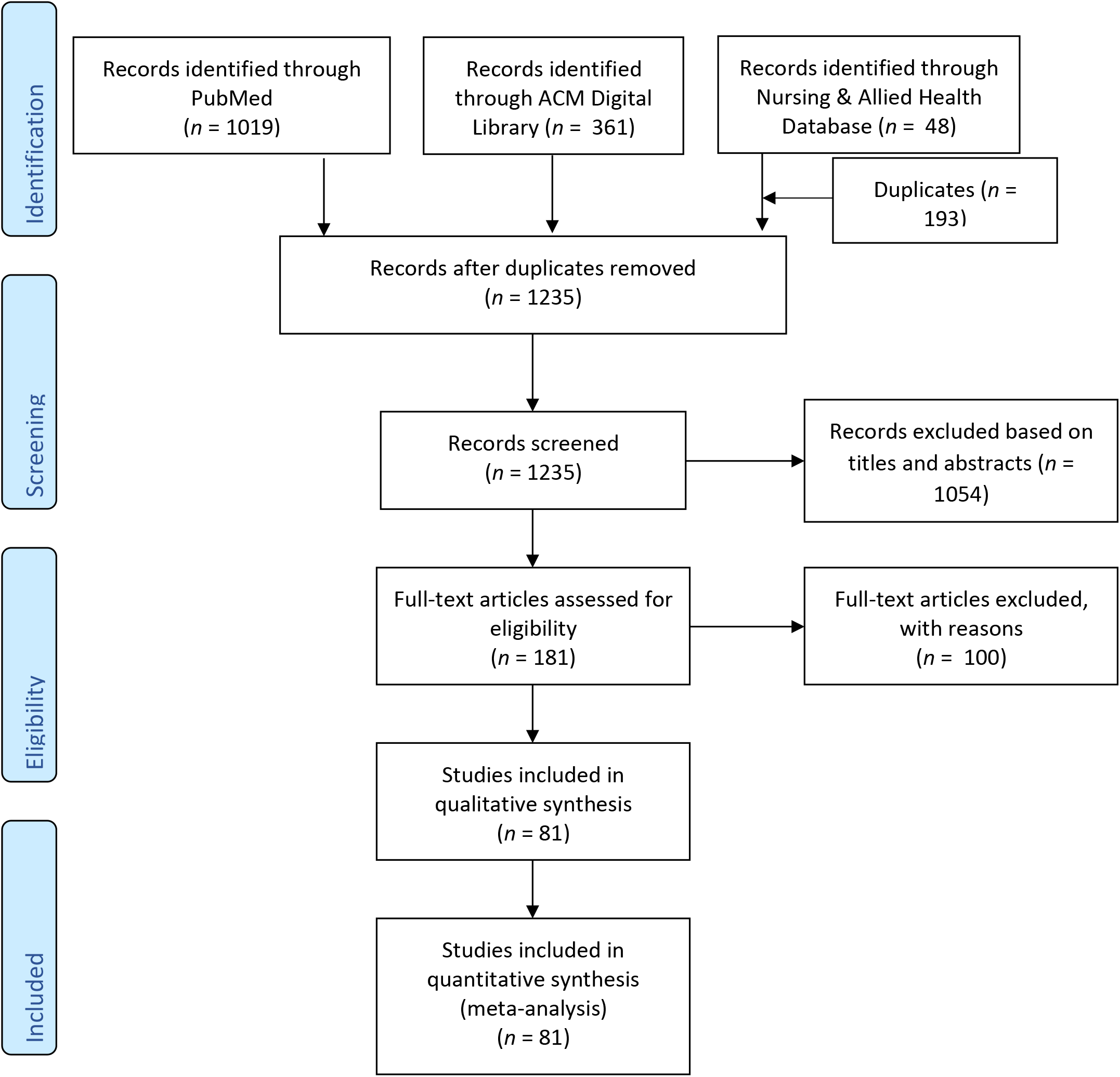
Flowchart of the literature search, screening, and selection

**Figure 3.**
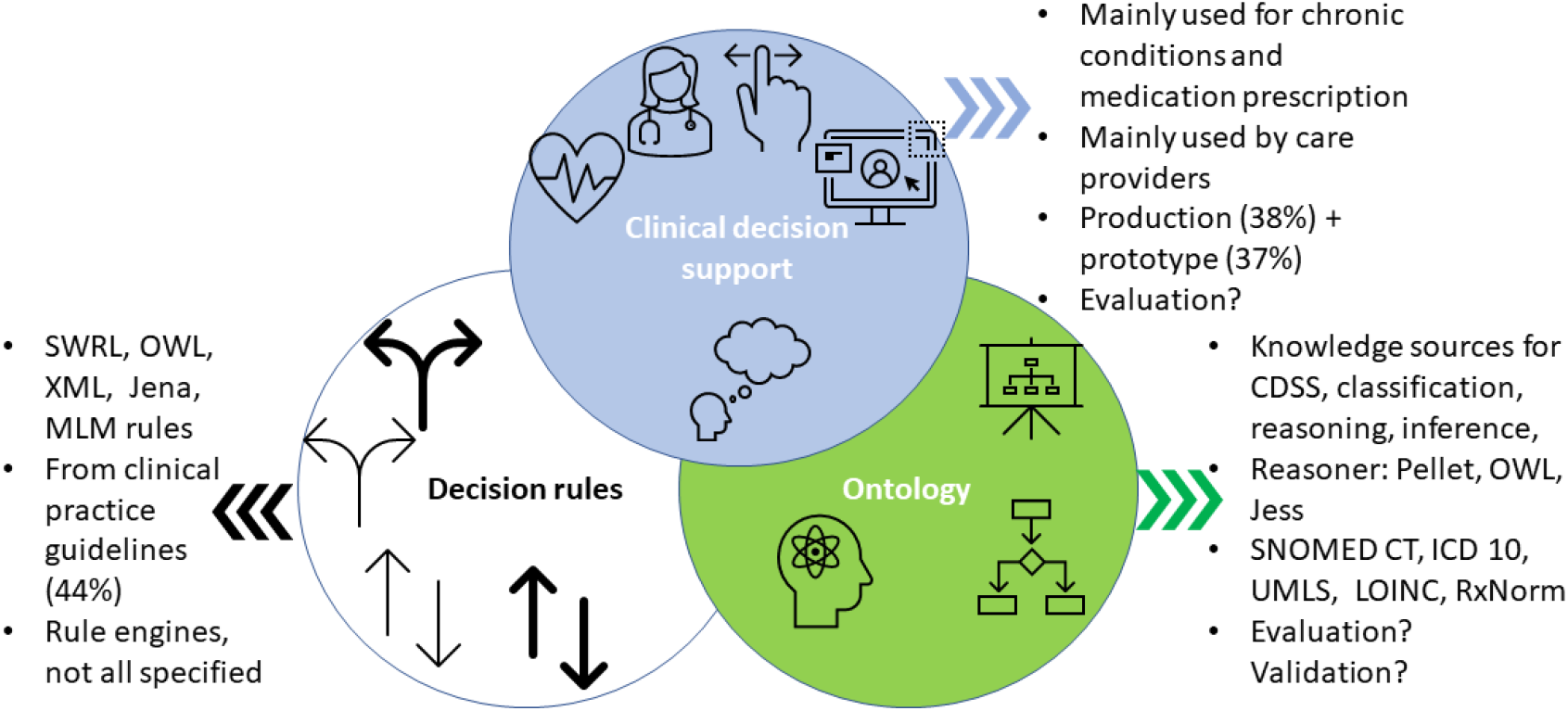
Initial knowledge framework on CDSS, CDSS rules, and ontology applications in CDSS

More than one-third (35.8%, 29/81) of CDSS was used for chronic condition management, prediction, or risk assessment. The chronic conditions included type 1 and 2 diabetes, hypertension, and asthma. Other significant domains included medication prescriptions (16%; 13/81), such as medication orders and detection of adverse drug events and drug-drug interactions, and cancer care (9.9%; 8/81). Most CDSS was designed and used by health care providers, and only 11.1% (9/81) were intended to be used by patients, some of which can be used by clinicians too. Most CDSS provide recommendations, suggestions, alerts, or reminders. Among all items we compared (Table 1), EHR evaluation had the least complete information. Some CDSS was implemented in production systems (38.3%; 31/81), excluding design, and some were implemented in prototypes (37%; 30/81), including experimental systems. Table 1 summarizes the essential characteristics of CDSS. We used the terms from the corresponding papers in all tables without changing them. For example, some papers used “physicians” as CDSS users, and others used “clinicians” as CDSS users. We used the authors’ terms in the tables.

**Table 1.** Basic CDSS profiles in included publications (*n* = 81)

| **Authors** | **Year** | **CDSS is used for**<br>**(Domain)** | **CDSS is used as** | **CDSS is used by** | **Architecture design**<br>**(production/prototype)** | **CDSS**<br>**evaluation** |
| --- | --- | --- | --- | --- | --- | --- |
| Abidi SS,<br>Manickam S<br>[44] | 2000 | The automatic<br>transformation<br>from XML to<br>clinical cases | Transformation tool | - | Architecture and<br>implementation | - |
| De Clercq PA,<br>Blom JA,<br>Hasman A, et<br>al [45] | 2000 | Drug interactions<br>or drug<br>contraindication in<br>ICU, family<br>medicine, and<br>psychiatry | Alerts | Physicians | Production | A hospital in the<br>Netherlands |
| Payne TH,<br>Savarino J,<br>Marshall R, et<br>al. [46] | 2000 | Medication errors<br>prevention and<br>detection | Alerts, reminders, notifications<br>to clinicians | Practitioners | In production, Veterans<br>Affairs hospital | - |
| Riesco AM,<br>Tomás RM,<br>Mira MJ [43] | 2000 | Therapeutic<br>decision analysis<br>in oncology | Recommendation of ideal<br>therapy | Physicians | - | - |
| Achour SL,<br>Dojat M, Rieux<br>C, et al [47] | 2001 | Safe blood<br>transfusion in<br>hospital settings | Alerts, consultation,<br>recommendations | Physicians | Prototype | 30 orders were<br>initially studied;<br>20 orders were<br>after<br>modifications |
| Séroussi B,<br>Bouaud J,<br>Dréau H, et al<br>[48] | 2001 | Prevention of<br>medication order<br>error in primary<br>care | Recommendations | General<br>practitioners | Prototype under<br>development | A simulated case |
| Karadimas HC,<br>Chailloleau C, | 2002 | Urology and<br>nephrology as<br>examples | Urgent or serious alerts | Physicians | Integrated into CIS and<br>patient record systems | Tested in urology<br>and nephrology<br>departments and |
| Hemery F, et al [49] |  |  |  |  |  | stand-alone used for education. |
| Ray HN, Boxwala AA, Anantraman V, et al [23] | 2002 | Diarrhea management | Patient assessment, treatment plan recommendations | Clinicians | CDSS integrated with a mini EMR | Plan |
| Das AK, Musen MA [50] | 2002 | Integration of time-related queries for clinical data | Integrated temporal query for clinical decision support | - | Software module | - |
| Barth C, Tobman M, Nätscher C, et al [24] | 2003 | Right diagnosis or treatment | Recommendations | Gastroenterologists | Prototype, web-accessible | Patient cases |
| Liaw ST, Sulaiman N, Pearce C, et al [52] | 2003 | Falls prevention | Recommend care plans | - | Prototype, Web API | Testing by clinicians and patients, and groups |
| Poon EG, Wang SJ, Gandhi TK, et al [51] | 2003 | To review and act on laboratory test results. | Manage abnormal results, generate letters to patients, send reminders for future tests | Outpatient physicians | Production | Pilot studies in two clinics with 20 physicians and nurses |
| Greenes RA, Sordo M, Zaccagnini D, et al [53] | 2004 | Centralized rule base and rule engine for CDS, e.g., medications, lab | Alerts, reminders | Physicians | Prototype | Case analysis |
| Ebrahiminia V, Riou C, Seroussi B, et al [54] | 2006 | Therapeutic plan for type 2 diabetes and hypertension | Recommendations for prescriptions | Clinicians | - | - |
| Kashyap V, Morales A, Hongsermeier T [55] | 2006 | Lipid management | Clinical recommendation or order | Clinicians | Production | Ontology inference performance |
| Abidi SR [57] | 2007 | Breast cancer follow-up care by PCP | Patient-specific recommendations | PCP | Production, two PCP clinics | - |
| Jannin P, Morandi X [58] | 2007 | Prediction of surgical positions in neurosurgery | Neurosurgeons | Neurosurgeons | Prototype | 159 surgical cases |
| Papageorgiou E, Stylios C, Groumpos P [60] | 2007 | Generic medical decision making support | Recommendations | Clinicians | Design | - |
| Stacey M, McGregor C, Tracy M [59] | 2007 | Neonatal intensive care | Early detection of sepsis, pneumothorax, and periventricular leukomalacia alerts | Clinicians | Prototype | Hypothetical case and hospital testing |
| Verlaene K, Joosen W, Verbaeten P [56] | 2007 | Generic design, two use cases: medication order and clinical pathway | Recommendations/suggestions | Clinicians | Prototype | Two scenarios |
| Cornalba C, Bellazzi RG, Bellazzi R [61] | 2008 | Risk management in hemodialysis | Suggestions for the risk profile of a patient and optimal decision | Clinicians, nephrologists | Design | Examples and case studies |
| Carenini M [65] | 2009 | Prediction, detection, monitoring of patient risk management | Alerts and notifications on efficient reaction or response to a risk situation | Clinicians | Ongoing design and ontology building | - |
| Dao TT, Marin F, Ho Ba Tho MC [63] | 2009 | Assessment, evaluation, treatment of clubfeet | Diagnosis, conservative treatment, and follow up | Patients and experts | Web-based system | Over 1000 cases used |
| Farion K, Michalowski W, Wilk S, et al [62] | 2009 | Acute pain in the emergency room, postoperative management of prostatectomy | Triage recommendations | Clinicians | Prototype | Feasibility |
| Zhou Q [64] | 2009 | Metabolism synthesis, diabetes, hypertension, hyperlipidemia, obesity, hyperuricemia | Recommendations | Clinicians | Experimental system | 20 clinical cases + 20 physicians |
| Basilakis J, Lovell NH, Redmond SJ, et al. [68] | 2010 | Telehealth, patient monitoring, health risk stratification to better manage the patient with chronic conditions | Alert, report, workflow modification | Clinicians | Framework, prototype | One case study |
| Borbolla D, Otero C, Lobach DF, et al [19] | 2010 | Preventive services, breast cancer screening | Recommendations, reminders | Physicians | Feasibility of Web service for SEBASTIAN incorporated with EMR in a hospital in Italy, production | Manually reviewed 210 patient cases |
| Lee J, Kim J, Cho I, et al [66] | 2010 | General design with a specific example in a laboratory context and hypertension | Alerts, recommendations | Clinicians | Prototype, testbed | 323445 test cases |
| Ongenaes F, Dhaene T, De Turck F, et al [67] | 2010 | ICU, sepsis, pattern detection in time series | Suggestions on time-related trends or notification of pathology classification | ICU clinicians | Framework | ICU use case |
| Wilk S, Michalowski W, Farion K, et al [69] | 2010 | Management of pediatric asthma exacerbation in the emergency department | Collect data, evaluate exacerbation severity, plan treatment | Emergency physicians | Simulated setting in hospital with HIS | Testing, 120 visits in 12 hours |
| Bouamrane MM, Rector A, Hurrell M [70] | 2011 | Preoperative risk assessment | Risk assessment, clinical recommendations | Clinicians | Software, mentioned implementation in practice | Example patient |
| Cao F, Sun X, Wang X, et al [72] | 2011 | Detect adverse drug events | ADE search and report | Clinicians | Production, web-based applications | - |
| Dao TT, Marin F, Bensahel H, et al [71] | 2011 | Assessment, evaluation, treatment of clubfeet, pediatric orthopedics | To generate a diagnosis, conservative treatment | Clinicians | Web application | Over 1000 cases |
| Lee CS, Wang MH [73] | 2011 | Diabetes diagnosis | Semantic description | - | Developed expert system, prototype | Experiments with cases |
| Bright TJ, Yoko Furuya E, Kuperman GJ, et al [75] | 2012 | Prescribing antibiotics | Alerts | Clinicians | Prototype | 81 patient records |
| Chniti A, Boussadi A, Degoulet P, et al [78] | 2012 | Medication orders | Suggestions on whether a prescription is valid | Clinicians | Prototype in hospital | Use case |
| Grando A, Farrish S, Boyd C, et al. [77] | 2012 | ADE reduction; safe and effective multi-drug treatment, COPD, type 2 diabetes, osteoporosis, hypertension | Recommendations | Clinicians with prescription rights | Prototype | Via evaluation scenarios |
| Koutkias V, Kilintzis V, Stalidis G, et al [76] | 2012 | Prevent adverse drug events | Alerts and recommendations | Clinical personnel | Knowledge framework | By experts and test cases; knowledge base authors testing the results via studies |
| Paterno MD, Goldberg HS, Simonaitis L, et al [79] | 2012 | Chronic condition management, diabetes, or coronary artery disease | Alerts, reminders | Clinicians | Production | Partners HealthCare, Wishard Memorial Hospital |
| Riaño D, Real F, López-Vallverdú JA, et al [74] | 2012 | Chronic comorbidities management | Personalized care plan recommendations | Healthcare professionals | Production systems in Italy, part of the K4CARE project | 916 patients were used to evaluate the CDSS |
| Artetxe A, Sanchez E, Toro C, et al [81] | 2013 | Diagnosis of Alzheimer's disease | - | Physicians | Production, Spanish research project | Ten patients |
| Corrigan D, Taweel A, Fahey T, et al. [84] | 2013 | Clinical prediction rules, Alvarado score | Clinical prediction | Clinicians | - | Three use cases |
| Farkash A, Timm JT, Waks Z [82] | 2013 | Hypertension | Recommendations, suggestions | Clinicians | Prototype | No user interface yet, two use cases for future |
| Haug PJ, Ferraro JP, Holmen J, et al [25] | 2013 | Pneumonia | Recommendation of diagnosis | Clinicians | An experimental system | Tested by sample patient data |
| Sáez C, Bresó A, Vicente J, et al. [83] | 2013 | Manage outpatient diabetes care in a telemedicine system | Recommendations for patient lifestyles on risks of diabetes | Physicians, patients | Production- telemedicine platform in Spanish | Ten patient records |
| Shojanoori R, Juric R [85] | 2013 | Remote patient monitoring system at care homes | Alarms, reminders, recommendations, or activating devices (e.g., heater) | Healthcare professionals | Experiment system | Preventive e-healthcare example |
| Wilk S, Michalowski | 2013 | Triage, diagnosis, therapeutic plan, disposition in the | Recommended actions, suggestions, patient data | Emergency department physicians | Pilot in production | 102 patients enrolled |
| W, O'Sullivan D, et al. [86] |  | emergency room; management of pediatric asthma exacerbations | management in the emergency department |  |  |  |
| Yao W, Kumar A [80] | 2013 | Generic clinical workflow, heart failure, diagnosis, and treatment | Recommendations | Healthcare professionals | Prototype | Example of patients |
| Yılmaz Ö, Erdur RC, Türksever M [87] | 2013 | Assistance in diagnosis and treatment | Recommendations | Physicians, dentists | Developed system based on SAMS | A dental scenario |
| Bau CT, Chen RC, Huang CY [88] | 2014 | Management of diabetes inpatients during surgery | Recommendations | Healthcare professionals | Prototype | 12 clinicians and 20 de-identified patients |
| Gallerani M, Pelizzola D, Pivanti M, et al [91] | 2014 | Laboratory repeated examination | Recommendation, warning on the appropriateness of the laboratory test | Physicians | Embedded in computerized physician order entry in two Italian hospitals, production | Three months of testing in two hospitals |
| Goldberg HS, Paterno MD, Rocha BH, et al [20] | 2014 | Chronic condition management, immunization schedules, pediatric head trauma | Recommendations, suggestions | Clinicians or patients | Production | Deployed at Partners HealthCare and trialed in multiple sites |
| Sesen MB, Peake MD, Banares-Alcantara R, et al [90] | 2014 | Lung cancer care, treatment selection | Recommendations | Physicians/experts | Prototype with a web interface | 4020 patient records retrospectively |
| Stewart SA, Abidi S, Parker L, et al [92] | 2014 | Pediatric cancer follow-up management | Recommendations | Patients | Embedded in an EHR portal in Canada | Evaluated by nurses and oncologists on content; domain experts evaluated rules |
| Wang HQ, Zhou TS, Tian LL, et al [89] | 2014 | A clinical pathway for deviated nasal septum | Provide more efficient and intelligent clinical procedures | Healthcare professionals | - | 224 patients with 11473 orders were selected to use/test |
| Delaney BC, Curcin V, Andreasson A, et al [94] | 2015 | Three use cases include diabetes, reflux symptoms, diagnostic support on abdominal pain, chest pain, and shortness of breath | Identifying patients with diabetes providing diagnostic support on effective medication for reflux symptoms. | Healthcare professional | Under development, the TRANSFoRm project | Three use cases and tests in a simulated environment |
| Jafarpour B, Abidi SR, Ahmad AM, et al [95] | 2015 | Alert environment avoid alert fatigue | Alerts | Clinicians | Framework | Five general practitioners |
| Robles-Bykbaeva V, López-Noresb M, Pazos-Ariasb J, et al [96] | 2015 | Speech-language therapy | Recommendations on therapy plans, generate content for students, monitor and evaluate patient progress | Speech and language pathologists, doctors, students, patients, relatives | In production, five institutions in Ecuador | 32 children assessed; 6 experts + 53 children evaluated |
| Shen Y, Colloc J, Jacquet-Andrieu A, et al [93] | 2015 | Optimal diagnosis, prognosis, treatment of gastric cancer | Suggestions on diagnosis, prognosis, treatment of gastric cancer | Clinicians and patients | - | Domain experts to provide feedback on CDSS; using 17 gastric cancer cases |
| Abidi SR, Cox J, Abusharekh A, et al [98] | 2016 | Safe prescription for patients with atrial fibrillation | Prescription eligibility check and authorization (Canadian), recommendations | Family physicians | Embedded within the IMPACT-AF ecosystem with a web-based stand-alone system | 100 de-identified patient cases |
| Goldberg HS, Paterno MD, Grundmeier RW, et al [100] | 2016 | Children with minor blunt head trauma | Recommendations | Physicians in the emergency department | One web accessed CDS service integrated with EHR in two sites, production | A multicenter trial |
| Marco-Ruiz L, Pedrinaci C, Maldonado JA, et al [101] | 2016 | Methods and models to enhance CDS service-oriented architecture as Linked Services | Alerts, recommendations | Clinicians | Models are complaint to Linked Data principles, semantic representation for CDSS services | The query of CDSS services |
| Wilk S, Kezadri-Hamiaz M, Rosu D, et al [97] | 2016 | Management of advanced chronic kidney disease | Identify correct physicians, handle exceptions, execute tasks, and change physicians. | Healthcare professionals, nephrologists, cardiologists, dieticians, nurse | Proof of concept implementation in OpenMRS | Simulated scenarios, fictitious patient data |
| Zhang YF, Tian Y, Zhou TS, et al [99] | 2016 | Inpatient management of type 2 diabetes | Recommendations for care plan | Clinicians | Prototype | 100 patient cases |
| Abidi S [106] | 2017 | Clinical management of comorbidities, including atrial fibrillation and chronic heart failure | Recommendations for safe and effective plans for comorbidity therapeutics | Family physicians | Web-based CDSS; stand-alone system | Domain experts on content; usability of CDSS by 10 PCP |
| Chen RC, Jiang HQ, Huang CY, et al [104] | 2017 | Diabetes management, outpatient | Recommendations on HbA1c target and medications | Endocrinologists or other physicians | Web system as a prototype | Ten patient records |
| Kopanitsa G [105] | 2017 | Treatment quality control as a use case | Check accuracy of mapping, transforming, and integrating data from multiple EHR | Clinicians | Production | 3436 treatment case records evaluated |
| Shang Y, Wang Y, Gou L, et al [103] | 2017 | Type 2 diabetes and hypertension in patient management | Suggestions for treatment decisions or lifestyles | Clinicians | Production, web services | Use cases |
| Zhang YF, Gou L, Zhou TS, et al [102] | 2017 | Chronic disease management, follow-up assessment | Recommended or suggested assessment/plan | Clinicians and patients | Deployed system, web service | Case study of patients with type 2 diabetes |
|  |  |  |  |  |  | (36162), follow-up assessments |
| Jin W, Kim DH [112] | 2018 | e-health services, sensors, device interoperability | Messaging | Healthcare professionals or patients | Prototype | Tested |
| Nakawala H, Ferrigno G, De Momi E [108] | 2018 | Surgical training system for thoracentesis | Surgical procedures planning, execution, practicing | Surgical trainees | Prototype | 10 participants in two experiments |
| Séroussi B, Guézennec G, Lamy JB, et al [109] | 2018 | Breast cancer management | Recommendations, action plans | Clinicians | Web API, in production | Case studies |
| Séroussi B, Lamy JB, Muro N, et al [111] | 2018 | Breast cancer management | Recommend best care plan for breast cancer patients | Clinicians | Web API, in production | Use cases |
| Shen Y, Yuan K, Chen D, et al [107] | 2018 | Infectious disease diagnosis, antibiotic prescription | Suggestions | Patients | - | Compare against other ontologies: DO, IDO, ROC for 215 infectious diseases; 84 records |
| Winter A, Stauber S, Ammon D, et al [110] | 2018 | Acute respiratory distress syndrome (ARDS) | Suggestions on mechanical ventilation; surveillance of ICU patients | ICU clinicians | Architecture framework | Use case for analysis |
| El-Sappagh S, Ali F, Hendawi A, et al [116] | 2019 | Monitor and manage type 1 diabetes | Real-time patient monitoring and care plan and lifestyle recommendation | Clinicians and patients | Cloud-based EHR system | Scenario |
| Jafarpour B, Raza Abidi S, Van Woensel W, et al [115] | 2019 | Safe and efficient comorbidity management | Recommendations | Clinicians | - | Local CIG integration engine test + surveys |
| Nguyen BP, Reese T, Decker S, et al [21] | 2019 | Detect potential drug-drug interactions | - | Clinicians | - | Two implementation use cases |
| Román-Villarán E, Pérez-Leon FP, Escobar-Rodriguez GA, et al [113] | 2019 | Manage cases of complex chronic illness | Recommendation and personalized treatment plans | Clinicians | Part of the PITeS-TiSS project, in the production system in Spain, DIRAYA | - |
| Semenov I, Osenev R, Gerasimov S, et al [114] | 2019 | Improve efficiency and efficacy of health care delivery | Recommendations, suggestions | Clinicians and patients | Production system connected with HIS | 15000 orders per day |
| Maldonado JA, Marcos M, Fernández-Breis JT, et al [117] | 2020 | Normalize clinical lab results; risk assessment for colorectal cancer screening | Support clinical data transformation to different formats; | Clinicians | Web-based platform | Two use cases with actual patients data |

Abbreviations: -, not specified; API: application programming interface; CDSS: clinical decision support systems; CIS: clinical information systems; COPD: chronic obstructive pulmonary disease; CPOE: computerized physician order entry; DIRAYA: the Electronic Health Record system within the Andalusian Public Healthcare System; DO: disease ontology; EMR: electronic medical records; EHR: electronic health records; ER: emergency rooms; HIS: hospital information system; HPO: human phenotype ontology; ICU: intensive care units; IDO: infectious disease ontology; KB: knowledge base; PCP: primary care providers; ROC: receiver operating characteristics; VA: Veterans Affairs.

### Results related to CDSS rules

Most CDSS rules were written in the languages of SWRL (11.1%; 9/81), Web Ontology Language (OWL; 13.6%, 11/81), extensive markup language (XML; 12.3%; 10/81), Jena rules (6.2%; 5/81), and medical logic module (MLM; 3.7%; 3/81). Two publications [109, 111] used N3 language, and two [82, 109] used the natural rule language (NRL). Table 2 details the CDSS rules for publications that can fill out three (except for the authors and publication years) or more cells in table 2 (*n* = 54).

**Table 2.** Comparison of CDSS rule characteristics in included publications (*n* = 54)

| Authors/<br>year | CDSS rule<br>language | CDSS rule<br>authoring/<br>editing | Rule sources | Rule<br>engine | Rule format/<br>management | Interoperability | Operation |
| --- | --- | --- | --- | --- | --- | --- | --- |
| De Clercq PA, Blom JA, Hasman A, et al, 2000 [45] | - | Knowledge Base Editor (KBE) | Guidelines | - | - | - | Patient data + events → guidelines ontology, models → recommended actions |
| Payne TH, Savarino J, Marshall R, et al., 2000 [46] | XML | - | - | - | Microsoft's Component Object Model (COM) | HL7 | Medication order + patient data → medication order monitor check → notifications |
| Achour SL, Dojat M, Rieux C, 2001 [47] | MLM | Newly presented tool | UMLS | Inference engine | Arden | - | Domain ontology → knowledge base → MLM + blood data + patient states |
| Karadimas HC, Chailloleau C, Hemery F, et al, 2002 [49] | MLM | The Arden/J framework developed by the authors | - | - | Arden | - | - |
| Ray HN, Boxwala AA, Anantraman V, et al, 2002 [23] | C | - | Guidelines from WHO | Guideline engine | CLIPS | - | Clinician enter data + EMR data + CLIPS rule → guideline engine + site configuration → recommendations |
| Barth C, Tobman M, Nätscher C, et al, 2003 [24] | Java | - | Textbook, multimedia | Inference engine | - | - | Patient data → inference engine + rules → case-based data + systematic data → recommendations |
| Liaw ST, Sulaiman N, Pearce C, et al, 2003 [52] | MLM, XML | - | Literature, physicians, and patients' input | - | Arden | HL7, XML | - |
| Kashyap V, Morales A, Hongsermeier T, 2006 [55] | OWL | - | Guidelines | ILOG, Cerebron (OWL ontology engine) | - | - | Patient data + event → rule engine + ontology engine → recommendations |
| Abidi SR, 2007 [57] | OWL | Rule authoring | Guidelines | GEM execution | The rule is specified in the | - | Computerize guideline → ontology → A family |
|  |  | model via an interface, self-developed |  | engine for CPG | logic element (class) of the CPG ontology |  | physician enters a patient's data via the CDSS interface, receives recommendations, explanation |
| Stacey M, McGregor C, Tracy M, 2007 [59] | XML | Jess | Data mining results and clinicians' input | - | - | - | Patient data + rules + ontology → alerts + archive |
| Verlaene K, Joosen W, Verbaeten P, 2007 [56] | XML | Eclipse Rich Client Platform, Graphical Editing Framework (GEF), ILOG JRules | Guidelines | Drools v2.4, Java and ILOG JRules v4.6 | - | HL7 RIM | Patient data + event + process → rule engine → recommendation |
| Cornalba C, Bellazzi RG, Bellazzi R, 2008 [61] | - | - | Guidelines, publications | Bayes network | Bayes rules | - | Patient data + monitoring → risk profile → Bayes network → optimal decisions |
| Carenini M, 2009 [65] | XML | Semantic Business Process Composer (SBPC) | Guidelines and data mining results | Business Process Execution Language (BPEL) | - | HL7 | - |
| Zhu Q, 2009 [64] | XML/SAGE | - | Guidelines | Re-inference | CLIPS rule set | - | Patient data + model → rules + inference → recommendations |
| Basilakis J, Lovell NH, Redmond SJ, et al., 2010 [68] | - | JBoss AS framework | - | JBoss Rules | jBPM (business process management) | HL7 | Clinical measures → rule engine + statistical analysis tool → recommendations |
| Borbolla D, Otero C, Lobach DF, | XML | - | Guidelines | - | SEBASTIAN rules | HL7 | Patient data + preventive service rules → reminder + explanation |
| et al, 2010 [19] |  |  |  |  |  |  |  |
| Lee J, Kim J, Cho I, et al, 2010 [66] | SAGE | uEngine, BRAIN | Guidelines | uEngine, BRAIN | - | - | Patient data + events + rule engine + workflow engine → alert or recommendation |
| Ongenaë F, Dhaene T, De Turck F, et al, 2010 [67] | SWRL | - | Galen ontology, medical databases | - | - | - | Patient data over time + ML + semantic reasoning → suggestions |
| Wilk S, Michalowski W, Farion K, et al, 2010 [69] | - | JADE (Java Agent DEvelopment Framework) | Literature, Cochrane library | - | - | Mirth Connect to enable communication between the MET3-AE and HIS; HL7 | - |
| Bouamrane MM, Rector A, Hurrell M, 2011 [70] | OWL | Protégé-OWL | Guidelines | JBoss Rules | - | - | Patient data + CDSS + ontology → risk assessment → recommendations |
| Cao F, Sun X, Wang X, et al, 2011 [72] | SPARQL queries, RDF | ADE ontology and knowledge base were used J2EE, DB2 V9, WebSphere V7. | Local hospital data, FDA documents, Structured Product Labeling (SPL), Linked Open Drug Data (LODD) | T-Box reasoner; A-Box reasoner | - | - | ADE knowledge + patient data → personalized ADE detection via semantic query, reasoning |
| Bright TJ, Yoko Furuya E, Kuperman GJ, et al, 2012 [75] | SWRL, queried by SQWRL | Protégé SWRL tab | Recommendation, assertive relational knowledge | Jess Rule Engine | Implemented in Java | - | Prescribing rules + ontology-driven alert module + patient database |
| Chniti A, Boussadi A, Degoulet P, | OWL | JRules, execute and manage business rules, | Business object model (BOM), the executable object model (XOM) | Jena | Java framework | - | OWL ontology/business object model entities + Jena → executable object model |
| et al, 2012 [78] |  | JRules OWL plug-in |  |  |  |  |  |
| Grando A, Farrish S, Boyd C, et al, 2012 [77] | SWRL, OWL | Protégé | Guidelines on COPD, diabetes mellitus, osteoporosis, and hypertension | Jess reasoner | Implemented in Java | Setting-independent rules | A repository of rules + polypharmacy treatment + ontology + patient data |
| Koutkias V, Kilintzis V, Stalidis G, et al, 2012 [76] | XML | A frame-based version of Protégé | Clinical guidelines and protocols | Inference engine: including CIG execution engine; Finite State Machine (FSM)-based execution engine | - | - | - |
| Artetxe A, Sanchez E, Toro C, et al, 2013 [81] | OWL-DL | - | Clinicians | - | - | - | Patient data + lab results → agent matchmaker → diagnosis assistance |
| Farkash A, Timm JT, Waks Z, 2013 [82] | The Natural Rule Language (NRL) | - | Clinical practice guidelines | - | - | HL7 CCD | Patient data in CDA → service, CDS rules, criteria → treatment recommendations |
| Sáez C, Bresó A, Vicente J, et al, 2013 [83] | Jess | HL7-CDA wrapper, developed by authors | Rules for physician assessment, rules for patient assessment, intermediate calculus rules, ADA recommendation rules, rule context information | Inference engine, pattern matching |  | HL7 – CDA input and output | CDA input → binding → inference → binding from rule results to CDA output → CDA output |
| Shojanoori R, Juric R, 2013 [85] | OWL/SWRL | - | Presented as an example, not in scale design | - | - | - | Patient data + monitoring data + contexts → ontology + reasoning → recommendations |
| Wilk S, Michalowski W, O'Sullivan D, et al, 2013 [86] | - | - | Requirements by emergency department physicians, guidelines | - | - | HL7 with HIS | Collect data (observation) → reasoning (decision) → action (recommendation) |
| Yao W, Kumar A, 2013 [80] | SWRL | Protégé + SWRLJessTab | Patient context, clinician context, resource context, location context, patient evaluation rule, patient diagnosis rule, patient treatment rule, prescription checking rule | Rule engine- Jess; Workflow engine- Drools-flow | Implemented in Java | - | Data from HIS + medical and context ontologies → rule + workflow engines → clinicians. |
| Yılmaz Ö, Erdur RC, Türksever M, 2013 [87] | SWRL | Protégé-OWL | Guidelines | Jess | - | - | Patient data + rule + knowledge → recommendation |
| Bau CT, Chen RC, Huang CY, 2014 [88] | Jena | Jena Rules | Disease, management, patient | Jena inference engine | OWL-DL, Protégé, Concept Explorer (ConExp), Jena rules | - | Patient data + patient ontology + Jena rules (from knowledge base) → IENA inference engine → recommendation |
| Gallerani M, Pelizzola D, Pivanti M, et al, 2014 [91] | XML | - | Publications, local domain experts | Drools rule engine | - | HL7 | User request → patient data + application server check → recommendations + archive |
| Goldberg HS, Paterno MD, Rocha BH, et al, 2014 [20] | XML | - | Guidelines | Operational decision manager (ODM) JBoss | - | HL7 CCD | Patient data + rules → recommendations |
| Sesen MB, Peake MD, Banares-Alcantara R, et al, 2014 [90] | OWL | - | NICE, BTS, ESMO, NCCN guidelines, Bayesian network from existing databases | - | Not specified | - | - |
| Stewart SA, Abidi S, Parker L, et al, 2014 [92] | SWRL | - | Recommendations | - | - | - | CPG computerization → CPG execution + patient profiles → personalized diary composition/ recommendation |
| Robles-Bykbaeva V, López-Noresb M, Pazos-Ariasb J, et al, 2015 [96] | Archetype | OpenEHR | Guidelines | - | - | - | - |
| Abidi SR, Cox J, Abusharekh A, et al, 2016 [98] | SWRL rules | - | Canadian clinical guidelines for atrial fibrillation | CPG execution engine | - | - | NOAC ontology + patient data → recommendation via auto-fill PDF |
| Goldberg HS, Paterno MD, Grundmeier RW, et al, 2016 [100] | XML | Operational Decision Manager (OPM) | Literature, prior trials | - | Production rules | HL7 | Patient data + prediction rules → recommendations |
| Zhang YF, Tian Y, Zhou | Jena | Protégé + Jena semantic framework | Clinical guidelines, care | Jena inference API | Jena rule | HL7 CDA as input and output | Patient data + ontology + rules → inference API → HL7 CDA → EMR |
| TS, et al, 2016 [99] |  |  | plans, domain experts, EMR |  |  |  |  |
| Abidi S, 2017 [106] | OWL-DL | Protégé | Clinical practice guidelines, domain experts; drug management protocols; expert-led reconciliation of CPGs | Reasoning engine | - | - | CPG ontology → computable CPG → comorbid CPG knowledge model → patient data + physician input → recommendation |
| Chen RC, Jiang HQ, Huang CY, et al, 2017 [104] | Jena rules, fuzzy rules | - | Guidelines, Technique for Order of Preference by Similarity to Ideal Solution (TOPSIS) | Jena inference engine | - | - | Patient data + fuzzy rules + drug knowledge base → recommendations |
| Shang Y, Wang Y, Gou L, et al, 2017 [103] | Jena | Rules are within the knowledge base, Protégé | CPG knowledge class, coding system class, patient class, disease, factors, treatment, recommendation | Jena reasoner + SPARQL | Java, Jena API, Jena rules | HL7 | Patient data + knowledge query + rules → rule engine → recommendation |
| Zhang YF, Gou L, Zhou TS, et al, 2017 [102] | Jena rules | Protégé | Clinical guidelines, nurses | - | - | HL7 and the Object Management Group (OMG) | Domain knowledge + patient data → assessment criteria → recommendation |
| Nakawala H, Ferrigno G, De Momi E, 2018 [108] | SWRL, OWL-DL | Protégé | Textbooks, interviews, web resources | - | - | - | Surgical procedures knowledge; instrument recognition |
| Séroussi B, Guézennec G, Lamy JB, et al, 2018 [109] | Formal module-driven rule language | - | PubMed, NCCN, ESMO, SEOM Clinical guidelines | - | N3 | FHIR to exchange input and output of CDS | DSS query FHIR server → +patient data → FHIR resources + rules + inference engine → recommendations |
|  | (NRL), N3, XML |  |  |  |  |  |  |
| Séroussi B, Lamy JB, Muro N, et al, 2018 [111] | N3 | - | Clinical guidelines, experience, prior cases | - | - | FHIR | Patient data → FHIR message → reasoning ontology → recommendations |
| El-Sappagh S, Ali F, Hendawi A, et al, 2019 [116] | SWRL | Protégé 5.1 editor | CPG, domain experts, web | Rule-based reasoner (e.g., Pellet) | Protégé, Jena API | Knowledgebase incorporated HL7 FHIR standards | Patient data + medical knowledge → reasoner → recommendation |
| Jafarpour B, Raza Abidi S, Van Woensel W, et al, 2019 [115] | OWL2 | - | Guidelines | Jena reasoner | - | - | Clinical state change + external events + rules + ontology → integration engine → recommendation |
| Nguyen BP, Reese T, Decker S, et al, 2019 [21] | CQL | - | Decision trees, guidelines | CQL engine | - | FHIR | Patient data + event → CQL library check → alerts or recommendations |
| Semenov I, Osenev R, Gerasimov S, et al, 2019 [114] | - | Rule editor has an interface, no name | Clinical guidelines | Rule engine and Bayes engine | - | FHIR JSON | - |
| Román-Villarán E, Pérez-Leon FP, Escobar-Rodriguez GA, et al, 2019 [113] | OWL | - | Clinical guidelines, healthcare professionals | - | OWL API | HL7 FHIR | Patient data + CPG + prior treatment data of the patient's → recommended treatment plan, personalized information |
| Maldonado JA, Marcos M, Fernández- | OWL, Archetype |  | Cancer screening protocols (USA, European) | Inference reasoning |  | Transforming, Semantic publishing, Mapping |  |
| Breis JT, et al [117] |  |  |  |  |  |  |  |

Abbreviations: -, not specified; ADA: American Diabetes Association; ADE: adverse drug events; AF: atrial affiliation; API: application programming interface; BTS: British Thoracic Society; CCD: continuity of care document; CDA: clinical document architecture; CDSS: clinical decision support system; CIG: computer interpretable guidelines; CLIPS: C Language Integrated Production System; COPD: chronic obstructive pulmonary disease; CPG: clinical practice guidelines; CQL: clinical quality language; DL: description logic; EMR: electronic medical records; ESMO: European Society for Medical Oncology; FDA: Food and Drug Administration; FHIR: Fast Healthcare Interoperability Resources; HIS: hospital information systems; HL7: health level 7; JESS: Jave Expert System Shell; MLM: medical logic module; NCCN: National Comprehensive Cancer Network; NICE: National Institute for Clinical Excellence; NOACs: New Oral Anticoagulants; NRL: natural rule language; OWL: W3C Web Ontology Language; RIM: reference information model; SPARQL: SPARQL protocol and RDF query language; SQWRL: a query language for OWL; SWRL: Semantic Web Rule Language; UMLS: unified medical language system; WHO: World Health Organization; XML: extensible markup language.

The most significant CDSS rule sources were from clinical practice guidelines (44.4%; 36/81). Other sources include domain expert input, publications (e.g., textbooks, papers), multimedia sources, and Internet resources. Two publications used data mining results as CDSS rule sources [59, 65]. CDSS rule authoring and editing tools were not routinely specified in the publications. Protégé [115] was the most popular tool to edit and author CDSS rules in publications with such detail. Several publications also describe developing such tools [49, 57, 83].

Rule engines are another technical detail that many publications did not specify. Among all publications with specified rule engines, Jena (7.4%; 6/81), inference engine (7.4%; 6/81), Jess (4.9%; 4/81), JBoss (3.7%; 3/81), guideline engine (3.7%; 3/81), Drools (2.5%; 2/81), and Bayes (2.5%; 2/81) were most often used. The column for CDSS rule operation in Table 2 summarizes how the CDSS rule works in a simplified manner. Many publications do not provide details on the working mechanism of CDSS rules within the system context.

Interoperability does not seem to be the focus of the publications. Only a few papers mentioned interoperability (Table 2). They all used HL7 CDA (Health Level 7, Clinical Document Architecture) or HL7 FHIR (Fast Healthcare Interoperability Resources) standards. However, we noticed that such measures are not specifically for CDSS rules but mainly for the input and output of CDSS.

### Results related to ontology

In the included publications, ontology is mainly used as a knowledge source for CDSS (39.5%; 32/81) and to facilitate classification (8.6%; 7/81), reasoning, and inference (7.4%; 6/81; e.g., identification recommendations or relationships). Ontologies were also used to specify CDSS rules (15%; 12/80) or provide general knowledge for the electronic medical record (EMR) or electronic health record (EHR) systems. In some cases, these two uses also overlapped (23.5%; 19/81; i.e., the ontologies were used to provide specified CDSS rules and general knowledge).

Many authors use reasoner and rule engines interchangeably. In this manuscript, we distinguish the two concepts in a workable manner to use them. We do not intend to define them universally and comprehensively. In our description, we use the term ***reasoner*** to refer to the inference for a consistency check or classification for an ontology, which can be part of an ontology tool or external. In our description, we use the term ***rule engine*** to refer to generating or providing recommendations by incorporating a patient’s data, contextual information, and medical knowledge (usually from an ontology or knowledge base) for CDSS. However, we used the authors’ choice of terms in tables without modification. Among the publications, the most common reasoners were Pellet (13.6%; 11/81), Jena (4.9%; 4/81), the OWL reasoner (3.7%; 3/81), Jess (2.5%; 2/81), and the Euler/EYE inference engine (2.5%; 2/81).

Ontology sources should include the content and code systems used to represent the content. Content may come from a popular textbook or clinical practice guideline. The content can be coded in a specific code system, such as SNOMED CT. Because the CDSS rule source has been specified in Table 2, we only included code systems that served as a source of ontology in Table 3; We used this format to avoid redundancy in the presentation. The most often used coding systems were SNOMED CT (11.1%; 9/81), ICD10 (the International Statistical Classification of Diseases and Related Health Problems, Tenth Revision; 4.9%; 4/81), Unified Medical Language System (UMLS; 4.9%; 4/81), Logical Observation Identifiers Names and Codes (LOINC; 3.7%; 3/81), RxNorm (2.5%; 2/81).

**Table 3.** Comparison of ontology roles in included publications (*n* = 36)

| **Authors/**<br>**year** | **Ontology is used**<br>**for** | **Specify**<br>**CDSS**<br>**rules/general**<br>**knowledge** | **Validated**<br>**by domain**<br>**experts** | **Reasoner** | **Ontology source/language** | **With CDSS** | **With**<br>**EMR/HIS** |
| --- | --- | --- | --- | --- | --- | --- | --- |
| De Clercq PA, Blom JA, Hasman A. et al, 2000 [45] | Knowledge source | Specify CDSS rules | Yes | - | - | - | - |
| Achour SL, Dojat M, Rieux C, 2001 [47] | Build MLM | Specify CDSS rules | Created by domain experts | Not specified | UMLS | Communicate | With prototype/HIS |
| Liaw ST, Sulaiman N, Pearce C, et al, 2003 [52] | - | - | Yes | - | ICD10-AM, ICPC2 | - | HL7 |
| Kashyap V, Morales A, Hongsermeier T, 2006 [55] | Classifier | Both | Yes, by knowledge base authors | OWL reasoner- Cerebra | Separate - ILOG | Communicate | Services on clinical data repository |
| Abidi SR, 2007 [57] | Knowledge source for recommendations | None specify/ Guidelines in GEM | Consult oncologist, individual clinical experience | - | Guidelines/OWL | GEM execution engine | - |
| Carenini M, 2009 [65] | Classify alerts and notifications | - | Experts validate taxonomy | - | JCAHO patient safety event taxonomy, ICPS, DOLCE | - | - |
| Farion K, Michalowski W, Wilk S, et al, 2009 [62] | Derive data, support, interface, configuration models | Both | Yes | - | - | Support ontology, executor | Implemented in prototype, web services |
| Ongenaë F, Dhaene T, De Turck F, et al, 2010 [67] | Classify temporal patterns | Both | Yes | DL reasoner-Pellet, probabilistic reasoner-Pronto | Rule-based reasoner | Decision module | New patients added to the ontology → automatically classified |
| Bouamrane MM, Rector A, Hurrell M, 2011 [70] | Classifications, recommendation | Both | Yes | Java-based Pellet | OPCS, ICD-10, NICE guideline/OWL | - | OWL, OWL API developed the system |
| Cao F, Sun X, Wang X, et al, 2011 [72] | Reasoning to get implicit relationships | Both | - | TBox and ABox reasoning over SNOMED | SNOMED-CT | Ontology mapping to ADE repository | Deployed in hospital as a Web application |
| Lee CS, Wang MH, 2011 [73] | - | Specify CDSS rules | Yes | - | Internet/OWL | - | - |
| Bright TJ, Yoko Furuya E, Kuperman GJ, et al, 2012 [75] | Generate prescribing alerts | Both | Yes | Jess rule engine | - | Alert module | Protégé + patient data + SWRL tab |
| Grando A, Farrish S, Boyd C, et al, 2012 [77] | Safe and effective polypharmacy prescription | Both | Yes | Jess reasoner | RxNorm, UMLS, COPD, hypertension, osteoporosis, diabetes guidelines | Conceptual test | Conceptual test with cases |
| Chniti A, Boussadi A, Degoulet P, et al, 2012 [78] | Author, execute, and manage business rules | Both | Yes | Java-based Jena | JRules OWL plug-in | Not integrated with HIS | CDS rules were implemented in HIS |
| Riaño D, Real F, López-Vallverdú JA, et al, 2012 [74] | Classification cases and treatment plans | Specify CDSS rules | Health care professionals provided some content | Jena | ICD10 CM, ATC/SDA modeling language | - | - |
| Artetxe A, Sanchez E, Toro C, et al, 2013 [81] | Infer clinical tests on patients to detect Alzheimer's disease | Both | Authored by domain experts | None specified; reasoning module | SNOMED CT, SWAN | The testing system of CDSS | Large scale Spanish research project |
| Sáez C, Bresó A, Vicente J, et al, 2013 [83] | Inference recommendation, assessment | Both | - | Rule-based reasoner | SNOMED CT | Case study test in a rule-based decision support module | Integrated with a telemedicine health platform |
| Yao W, Kumar A, 2013 [80] | Infer context information, patient conditions, rules for CDSS | Both | - | Pellet reasoner plug-in for Protégé 3.4 | Jess rule engine, workflow engine-Drools-Flow 5.2 | Scenario test, integration with clinical workflows | CONFlexFlow implementation, web-based services |
| Yılmaz Ö, Erdur RC, Türksever M, 2013 [87] | Inference new facts | Both | - | - | ICD/OWL | - | - |
| Bau CT, Chen RC, Huang CY, 2014 [88] | Infer recommendations for patients | Both | Yes | Jena | Jena inference engine | CDSS prototype | Patient data + CDSS |
| Sesen MB, Peake MD, Banares-Alcantara R, 2014 [90] | Ontological guideline rule inference | Specify CDSS rules | - | FaCT++ | NICE Guidelines, OWL, mapping to SNOMED CT | OWL API + FaCT ++ semantic reasoning + probabilistic reasoning | JDBC connect to patient records database |
| Stewart SA, Abidi S, Parker L, et al, 2014 [92] | Computerized CPG | Specify CDSS rules | Domain experts evaluate rules | Pellet reasoner | CPG, OWL-DL | - | Embedded in an EHR portal in Canada |
| Abidi SR, Cox J, Abusharekh A, et al, 2016 [98] | Knowledge sources, criteria | Specify CDSS rules |  | OWL reasoning | AF clinical practice guidelines/OWL | Computerized NOAC authorization decision support system | Can be integrated with EHR or stand-alone web version |
| Marco-Ruiz L, Pedrinaci C, Maldonado JA, et al, 2016 [101] | Describing CDS services, functional, nonfunctional, and data semantics | Both | Use case validation | OWL Horst | SNOMED CT, Dublin Core/OWL | - | - |
| Zhang YF, Tian Y, Zhou TS, et al, 2016 [99] | Infer diagnosis, recommendations | Both | Yes /examined by domain experts | Pellet | An inference API –Jena rules | Experiments on CDSS | CDA format used to convert between EMR and CDSS |
| Abidi S, 2017 [106] | Knowledge sources | Specify CDSS rules | Yes, via evaluation by independent cardiologists, PCP | Pellet | Practice guidelines, domain experts; protocols | Communicate | Stand-alone EMR |
| Shang Y, Wang Y, Gou L, et al, 2017 [103] | Infer personalized recommendations | Both | Yes | None specify | Jena API | Web services for CDSS | Web API by using patient data + CDSS |
| Zhang YF, Gou L, Zhou TS, et al, 2017 [102] | - | - | - | Pellet | SNOMED CT /OWL-DL | - | EMR and other sources |
| Nakawala H, Ferrigno G, De Momi E, 2018 [108] | Knowledge sources | Specify production rules | Yes | Pellet | Journal article, physician, online resources, HONcode search engine; FMA, IAO, BFO, W3C time ontology | - | - |
| Séroussi B, Guézennec G, Lamy JB, et al, 2018 [109] | Knowledge sources | Specify CDSS rules | - | Euler/EYE inference engine | NCI Thesaurus, LOINC, SNOMED CT, OWL | - | - |
| Séroussi B, Lamy JB, Muro N, et al, 2018 [111] | Knowledge sources | Specify CDSS rules | - | Euler/EYE inference engine | Guidelines | Web services on CDSS | - |
| Shen Y, Yuan K, Chen D, et al, 2018 [107] | Identify disease and recommend treatment | Both | - | - | IDO, NCBI Taxonomy, HPO, DrugBank, DO, Internet, Guidelines | Clinical decision module | Patient data + ontology + clinical decision module + user interaction module |
| El-Sappagh S, Ali F, Hendawi A, et al, 2019 [116] | Provide a personalized care plan | Both | Yes | Pellet | SWRL rule engine, Jena API | Cloud-based CDSS | FHIR patient data + CDSS + ontology → FHIR → EHR |
| Jafarpour B, Raza Abidi S, Van Woensel W, et al, 2019 [115] | Provide domain knowledge and integration policies | - | A medical expert validated the integration solution; | Jena reasoner | SNOMED, Drug Bank/ OWL | - | Integration framework with web API, CIG server and |
|  |  |  | medical experts instantiate integration policies |  |  |  | client, client connect POC |
| Román-Villarán E, Pérez-Leon FP, Escobar-Rodriguez GA, et al, 2019 [113] | - | Specify CDSS Rules | Yes, by clinical researchers | - | Mapped to SNOMED CT | - | - |
| Maldonado JA, Marcos M, Fernández-Breis JT, et al [117] | Classification of risk level | Both | - | OWL reasoner | LOINC | Web-based platform | Patient data from EHR + platform |

Many included publications also did not include complete information for ontology validation. Approximately 20 publications mentioned some validation, including validation or evaluation by domain experts (24.7%; 20/81). Some ontologies were authored by domain experts [47, 55]. Table 3 shows additional details on the roles of ontology in the publications, although Table 3 includes only the publications that can fill out three (except for the authors and publication years) or more cells (*n* = 36).

### Other results

After cleaning, discussion, and consolidation, we had 30 code groups and 221 final codes used in the ATLAS.ti (Appendix 2). These codes and code groups guided our results analysis and synthesis; these codes and code groups also helped us form the starting structure for the ontology we are building [41], i.e., the ontology for CDSS rule management and maintenance. Figure 4 illustrates the clinical domains of CDSS within included publications. Figure 5 shows a word cloud image generated from ATLAS.ti based on the codes used in all included publications. In the publications, we also found many large international collaborations [94, 101, 110] and solo author publications [57, 64, 105, 106]. We also noticed that some publications were missing necessary information for explaining the mechanisms of the systems that can be critical for reproducibility. Some examples include the architecture diagrams of CDSS, CDSS rule engine, CDSS rule language, how CDSS rules are managed at the backend, and integration mechanisms among CDSS rules, ontology, and EHR/EMR systems.

**Figure 4.**
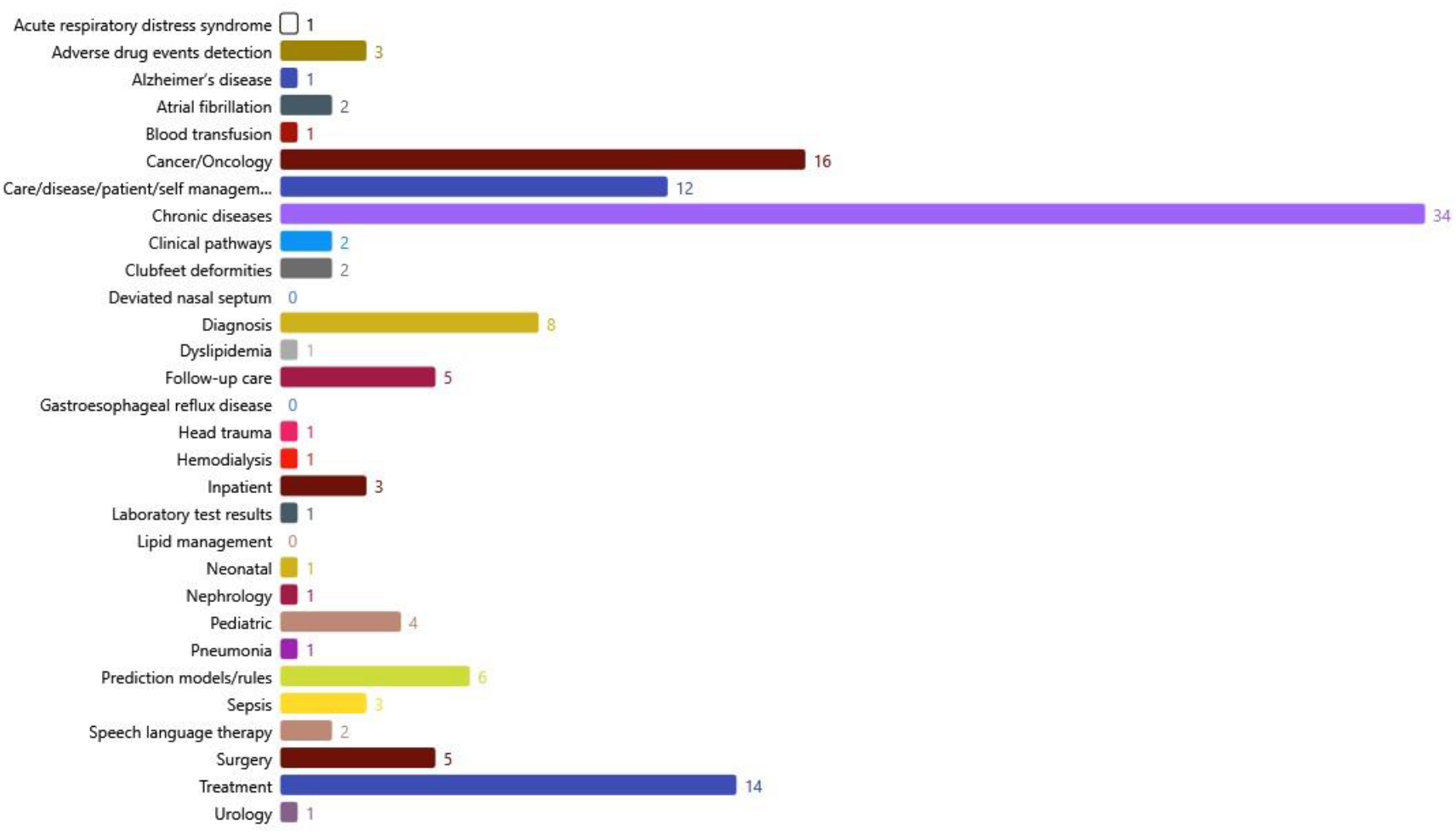
Bar chart generated from ATLAS.ti showing the clinical domains of CDSS in included publications.

**Figure 5.**
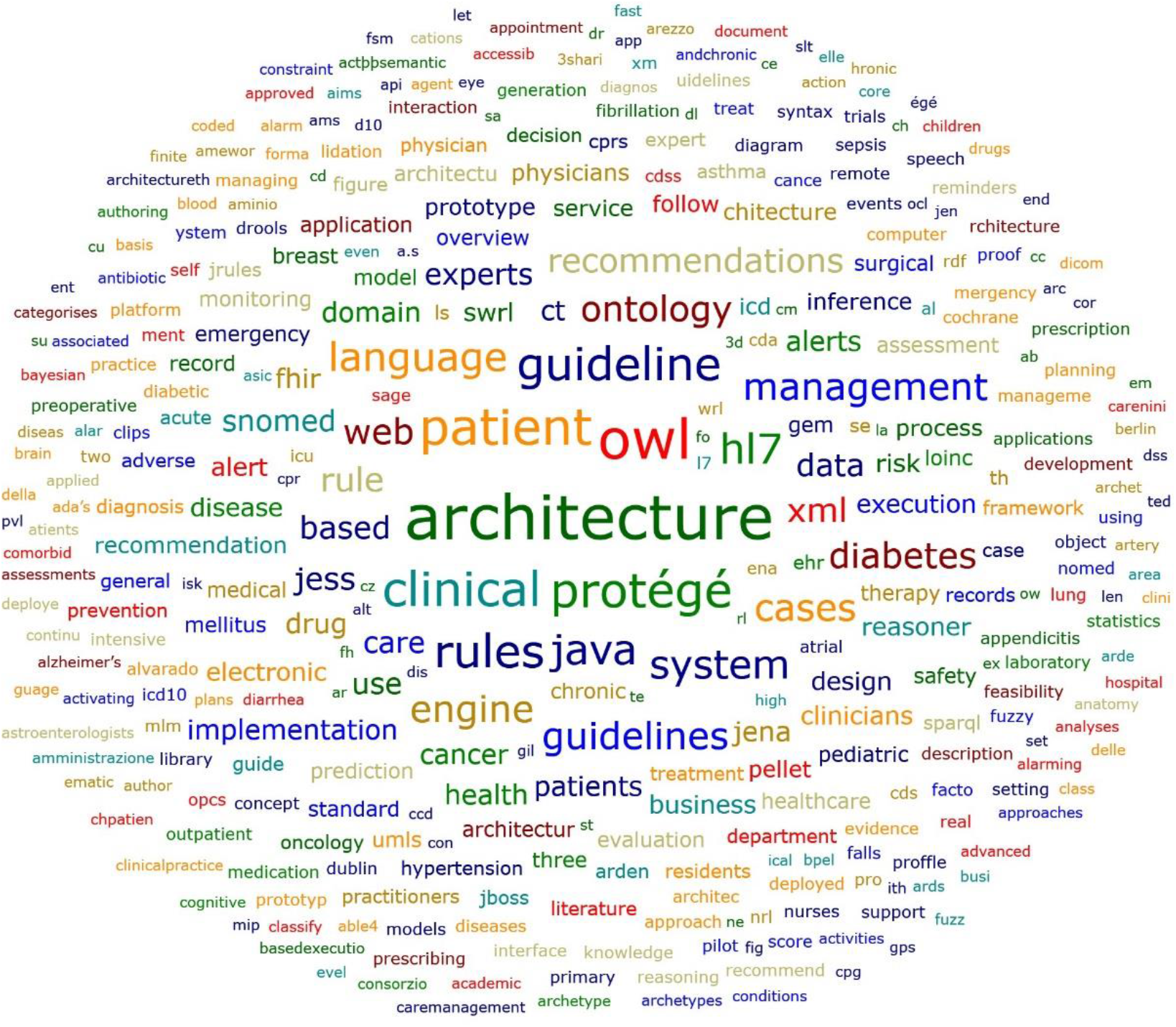
Word cloud generated from ATLAS.ti based on codes in the included publications

Based on our review, PubMed is still the clear dominant source to supply publications in this field. Approximately 90.1% (73/81) of publications were identified via PubMed. The ACM Library added only eight publications after removing duplicates.

We have followed the PRISMA 2020 checklist in writing this systematic review. However, we realize that PRISMA 2020 is intended to be used by outcome-oriented studies, which is not the focus of our systematic literature review. The publications included in our review are about design, development, and implementation. Therefore effect measures or certainty assessment were irrelevant items. We reported 18 items (out of 27 items, Appendix 4) in the full-text paper and ten in the abstract (out of 12 items, Appendix 5).

## Discussion

### Summary of results

Although CDSS has proved its effectiveness in clinical care for decades, the reusing and sharing of machine-processable CDSS rules has not been achieved. Ontology has the potential to facilitate interoperable CDSS rules. We conducted this systematic review on CDSS rules, ontology, and its applications in CDSS. Our systematic review shows the broad clinical application domains of CDSS in chronic condition management, medication order, and cancer care. Most CDSS is designed for health care providers, and most of the CDSS rules are based on clinical practice guidelines. Although we found details about CDSS rules, we noticed inconsistent presentations or unavailable information on CDSS rule language, CDSS rule engine, and evaluation details. We also observed inconsistent technical details related to ontology purposes, reasoners, mechanisms of connection, or communication with CDSS or EMR/EHR/HIS. Although some publications describe such a level of detail, not all publications include this information. It can be hard to perform reproducibility and further improvement without such information.

Although reusing and sharing CDSS artifacts is a challenge with very good recognition [1, 101], the reusability, customization, and shareability of CDSS rules are not yet a common focus, not even in publications that focus on CDSS rule editing [35, 118, 119]. We feel these topics are important, especially using ontology to achieve the goals. Therefore we conducted this literature review. Marco-Ruiz et al. [101] showed how to use CDSS artifacts in the Linked Data framework [120] by leveraging Semantic Web technologies, especially ontology. However, that work was at a higher level to describe the concepts without tangible tools implemented in clinical practices. Our motivation to build a CDSS ontology [41] aligns well with their vision. We are trying to shorten the gap between the high-level vision on these realistic issues (reusing and sharing CDSS artifacts) [1, 101] and CDSS operations in the clinical world by appraising existing efforts and using a more generic approach. The first step in filling this gap is constructing an upper-level CDSS ontology.

### Significance of the work

We systematically reviewed literature about ontology applied in CDSS. Our approach illustrated the state-of-the-art applications of ontology in CDSS rule management. These applications present excellent potential in reusing and sharing CDSS artifacts, which are critical to clinical care quality and patient safety. Currently, it is very costly to create, maintain, and reuse these rules. Although some authors recognized this benefit [1, 35, 101], none have conducted a systematic review. Our literature review thoroughly analyzed the topic, established the knowledge framework, and created a comprehensive collection that can inform efforts to design or improve future CDSS, create a tutorial on the subject for newcomers in the field, and set the foundation for the CDSS ontology we are building.

We also recommend that authors include essential technical details in publications focusing on this topic. These details include CDSS application domain, CDSS users, CDSS notification types, CDSS evaluation (what, how, by whom), CDSS rule sources, CDSS rule language, CDSS rule engine, CDSS operation mechanism, ontology use purposes, ontology sources (both content and code systems), ontology validation, reasoner, connection/communication mechanism with CDSS, and EMR/EHR. We encourage authors to include such details to help readers reference, compare, and increase the reproducibility of the reported work.

### Interpretation of the results

Rule engines are critical components of CDSS. The rule engine executes rules and patient data and context information to produce a result, e.g., an alert or a recommendation [1]. Jess is a rule engine and development environment in Java [121]. We noticed in our review that Jess is one of the famous rule engines used repeatedly. Jess can be used to develop rule-based expert systems or create Jess rules. In the popular tool Protégé, the plug-in API SWRLJessTab can convert SWRL rules to Jess rules. Jess rules can be used by Jess Rule Engine, which has many implementations in rule-based expert systems [121]. In addition to Jess, Jena and Drools were used repeatedly among the included publications. Jena is an API in Java designed to support rule-based inference and use RDF (Resource Description Framework) graphs [122]. Jena.java API can handle OWL models and is a popular framework for managing RDF/OWL descriptions [88]. Drools is a business rule management system, including a rule engine [123]. Drools also have SWRL API, which can support SWRL and SQWRL. SQWRL can query SWRL.

Reasoning via a reasoner is a critical characteristic for many ontologies, even though the current reasoning is still in the first-degree logic. Reasoning can be used for three main functions: consistency check, classification, and realization [124]. Several publications included in this systematic review specified the classification roles of the ontology and the reasoners (listed in Table 3). The OWL group has curated an updated list of OWL reasoners at the University of Manchester in the UK [124]. Parsia and colleagues compiled and compared the current OWL reasoners and their performances via the competition report [124]. Here, we only briefly discuss the reasoners used in the included publications.

Among the included publications, both Pellet and Jena are popular reasoners (Table 3), and other used reasoners include FaCT++ [90], Z3 Solver reasoner [97], Euler/EYE inference engine [109, 111], OWL Horst [101], and OWL Cerebra [55]. Among these reasoners, Pellet [125] is Java-based, and it can work on SWRL rules and ontologies written in OWL2. SWRL was initially designed as a rule language for Semantic Web technologies [126]. A user needs the rule language and an editor (e.g., Protégé SWRL tab) to write, revise, and query the rules. SWRL can be queried by SQWRL (a query language for OWL) or SPARQL (SPARQL Protocol and RDF query language). Then reasoners can be used to conduct reasoning based on the rules and facts defined in the ontology or knowledge base. Protégé-OWL [127] provides an editor for SWRL rules. Protégé SWRL editor is another example.

Not all ontologies or knowledge bases were formally evaluated, validated, or authored by domain experts. Only 25% of publications mentioned about evaluated or validated by domain experts. A formal assessment or validation is critical, regardless of a manually built ontology (or knowledge base). For some ontologies (or knowledge bases) derived from other automatic methods (e.g., machine learning algorithms), the validation is even more critical to ensure the validity of the results from automated processes. Testing has not been conducted consistently across the publications. Some have been formally tested with patient databases in the production systems, some were tested via prototypes, and some were tested conceptually. Technically, all such testing is valuable, to some extent. Without some types of testing, the validity of the work can be questionable. Some ontologies were authored by domain experts, which provides greater validity than those involving nondomain experts while constructing the ontology.

Also, not all publications clearly state whether the evaluation was conducted on the CDSS. For those publications with CDSS evaluation, not all provide adequate details on what was evaluated, how the evaluation was completed, and who the evaluators were.

We noticed that not all included publications provided the needed technical details during the literature review, such as the CDSS rule engine, rule language, and ontology reasoner. Some people may argue that such information can vary dramatically during implementation. However, we believe that including such details will present the work more robustly, help users better understand the system’s mechanism, provide possibilities for others to reproduce the work, and enable others to compare the technical solutions and design more optimal options in the future.

One systematic review by Dissanayake and colleagues [40] was similar to our review. However, our review has several distinct characteristics that complement the literature with unique insights about CDSS rules, ontology, and ontology applications, especially in rule management. For example, we focused on the mechanism of these characteristics in the HIS/EHR/EMR systems. In contrast, Dissanayake’s review focused on the reasoning processes during clinical decision-making, especially in forming the ontologies to support complex cognitive processes. Their study also compared evaluation metrics and evidence generated among the included publications, whereas we focused on the CDSS rule authoring, management, implementation, and ontology validation.

Our coding revealed factual evidence without any interpretations, which differs from qualitative data analysis and coding. Our discussions focused on the scope of coding, definitions of codes, the granularity of coding, dimensions, organization, categorization, and combinations of codes, articulating these points and obtaining consensus.

### Knowledge gap and challenges moving forward

Although the importance and challenging nature of reusability and sharability of CDSS artifacts have been well recognized and documented, various efforts are focusing on this topic because of the potential of the Semantic Web technologies and ontology [1, 101]. However, the steps are mainly at the conceptual level without tangible tools to implement in the real world. No CDSS ontology bridges the high-level thinking, planning presented by different groups, and real-world implementations and operations of the process. This knowledge gap motivated our efforts to build a CDSS ontology that focuses on CDSS rule management and maintenance [41]. Moving forward, one challenge is more on the marketing side: let CDSS artifacts developers, vendors, and healthcare professionals, i.e., end-users, be aware of the CDSS ontology and adopt it in their work.

### Strengths

This systematic review focuses on CDSS rules and ontology roles in CDSS and mechanisms of CDSS in clinical practices or prototypes. We established the initial knowledge framework on the intersections of CDSS, CDSS rules, and ontology. We summarized detailed information regarding the reasoner, rule engines, ontology, and CDSS rule formats used in each included publication. We also provided summaries (Tables 1–3) of valuable references for designing or improving systems. The side-by-side comparison also gives structured guidance for preparing future publications. We hope the knowledge framework, detailed summarizations, and comparisons can guide and enlighten future improvements and designs more explicitly in tangible ways.

### Limitations

We recognize that our review has limitations. We were unable to include publications written in a language other than English. Also, we only included publications with full-text available. We had to exclude one thesis [128], which was a search result from the ACM Library, due to the unavailability of the full-text publication. Also, we excluded some publications that focused on the CDSS rule [35, 118, 119]. Although such publications had a similar focus to one aspect of our systematic review, none specified an ontology component in the publications. We also noticed that the publications on CDSS rule authoring and managing tools are from Partners HealthCare/Harvard Medical School.

We also exclude another category of papers that discussed best practices in CDSS rule management, such as Wright A et al. [10]. Although such publications can inform CDSS design, testing, and operation processes, they did not meet the focused criteria for this systematic review.

Interestingly, our two papers [129, 130] were not identified via our literature search strategies. The main reason such a search is missing is the keywords used in these papers. CDSS was not specified as a keyword in the two papers, although the content was undoubtedly within the scope of this review.

Unfortunately, these two publications may not be the only papers missed by this literature search. This challenge is common to how our current literature databases are organized and how we conduct a literature search. In the future, this challenge can be minimized by carefully choosing all keywords included in the publications to maximize the possibilities found during the literature search. This is also a lesson for all authors.

## Conclusions

The reuse, management, and maintenance of CDSS rules are critical and challenging for clinical care. Ontology could facilitate the reusing and sharing of CDSS artifacts, e.g., CDSS rules. However, there is no CDSS ontology yet, and we are building one. We attempt to bridge the high-level visions and operational level efforts via the CDSS ontology to bring tangible applications to the field. Current publications present incomplete technical details on CDSS rules and ontology. We recommend future publications include more detailed information about the architecture diagrams, mechanism of connection among ontology, CDSS rules, EHR/EMR, CDSS rule language, reasoner, the rule engine, validation or authorization of ontology and CDSS rules, the purpose of the ontology, ontology source, and the management and maintenance of CDSS rules. Such details will increase the study’s reproducibility and help readers optimize their designs and development. In addition, we hope the established knowledge framework and summarizations of the included publications can guide future improvements and designs more explicitly.

## Data Availability

All data produced in the present study are available upon reasonable request to the authors

## Acknowledgment

This work is supported by the National Institute of General Medical Sciences of the National Institutes of Health under Award No. R01GM138589 and partially under P20 GM121342. We thank Professor James Cimino for his valuable feedback and suggestions on the manuscript.

## Competing interests

None to disclose.

## Appendices

**Appendix 1:** Initial draft of codes and code groups used in reviewing/coding

**Appendix 2:** Refined version of codes and code groups used in reviewing/coding

**Appendix 3:** All included paper lists (*n* = 81) in this systematic literature review

**Appendix 4:** PRISMA 2020 item checklist reported in our systematic literature review

**Appendix 5:** PRISMA 2020 for Abstracts checklist reported in our systematic literature review

## Notes

### Competing Interest Statement

The authors have declared no competing interest.

