## Appendix 1 for "A systematic review of ontology-based clinical decision support system rules: usage, management, and interoperability"

| name | comment | codegroup 1 | codegroup 2 |
|---|---|---|---|
| Architecture diagram | | Architeture design | |
| Author | Maybe we can start from the family name of the first author to record each paper. | | |
| BFO | | Ontology Sources | |
| Clinicians | Can be the evaluators | Evaluation | |
| DO | | Ontology Sources | |
| Drools | Workflow engine-Drools-flow | Rule Engine | |
| DrugBank | | Ontology Sources | |
| Expert | | Ontology Sources | |
| Feasibility | | Evaluation | |
| Galen Ontology | | Ontology Sources | |
| GEM execution engine | | Rule Engine | |
| Guidelines | | Ontology Sources | |
| HPO | | Ontology Sources | |
| ICD-10 | | Ontology Sources | |
| IDO | | Ontology Sources | |
| Inference engine | | Rule Engine | |
| JENA inference engine | Jena reasoner | Rule Engine | |
| JENA rule engine | | Rule Engine | |
| JENA rules | | CDSS Rule Format | |
| Jess reasoner | | Rule Engine | |
| Jess Rule Engine | | Rule Engine | |
| Jess rules | | CDSS Rule Format | |
| JRules OWL plug-in | | Rule Management | |
| Literature | | Ontology Sources | |
| LOINC | | Ontology Sources | |
| Medical records | Also can be patient records, patient cases, can be identified or de-identified | Ontology Sources | Evaluation |
| NCBI taxonomy | | Ontology Sources | |
| NCI Terminology | | Ontology Sources | |
| Ontology inference performa | | Evaluation | |

| | | | |
|---|---|---|---|
| OWL | | CDSS Rule Format | Ontology Language |
| Pattern matching | ? | Rule Engine | |
| Production system | | Architeture design | |
| Protege | It can be used as a visual editing tool for OWL rules | Rule Management | |
| Prototype | | Architeture design | |
| Protégé SWRL tab for editing | | Rule Management | |
| Protégé+ SWRL Jess Tab | | Rule Management | |
| Publication year | This is also used to record the paper. | | |
| Reasoner: COSI (Closed Worl | | Rule Engine | |
| Rule authoring interface | | Rule Management | |
| Rule-based reasoner | (e.g., Pellet) | Rule Engine | |
| SNOMED CT | | Ontology Sources | |
| SWAN | | Ontology Sources | |
| SWRL rules | | CDSS Rule Format | |
| UMLS | | Ontology Sources | |
| Use cases | Can be ICU or other units; also called scenarios. | Evaluation | |
| Web resources | | Ontology Sources | |
