## Appendix 2 for "A systematic review of ontology-based clinical decision support system rules: usage, management, and interoperability"

| Codebook-literature review | | |
|---|---|---|
| **Code** | **Comment** | **Code Groups** |
| Action variables | | |
| activating devices | | |
| Acute pain | | |
| adverse drug event | | |
| Adverse drug events detection | | CDSS Purpose-Domain<br>Ontology Purpose<br>CDSS Purpose-Type |
| alarm | | |
| Alerts | Can be drug-drug interactions, or drug-dosage or other types of alerts | Ontology Purpose<br>CDSS Purpose-Type |
| Alzheimer's disease (AD) | | CDSS Purpose-Domain |
| Antibiotic prescribing | | CDSS Purpose-Domain<br>Ontology Purpose<br>CDSS Purpose-Type |
| Archetypes | | |
| Architecture diagram | | Architeture Design |
| Arden syntax | | CDSS Rule Format |
| Asthma | | |
| Atrial fibrillation | | |
| Author | Maybe we can start from the family name of the first author to record each paper. | |
| Bayes engine | | |
| BFO | | Ontology Sources |
| Blood transfusion | | |
| BRAIN rule engine | | |
| Breast cancer | | CDSS Purpose-Domain |
| Business Process Execution Language (BPEL) | | |
| business logic | | |
| business rules engine | | |
| Business rules-ILOG | | Rule Management<br>CDSS Rule Format |
| C++ | | |
| Cancer | | |
| Care management | Including patient management, disease management | CDSS Purpose-Domain<br>Ontology Purpose |
| CCD | | |
| CDA | | |

| | | |
|---|---|---|
| Chronic diseases | | |
| Chronic heart failure | | |
| Chronic kidney disease | | |
| Classifier | Including to classify temporal patterns, to categorize patients into groups | Ontology Purpose |
| clinical pathways | | |
| Clinicians | Can be the evaluators | CDSS Rule Sources<br>CDSS Evaluation (who, what, how, where) |
| CLIPS ruler sets | | |
| Clubfeet deformities | | |
| consultation document | | |
| COPD | | |
| Coronary artery disease | | |
| CPG rule syntax | | |
| de facto standard | | |
| Decision variables | | |
| Diabetes | | CDSS Purpose-Domain |
| Diagnosis | It ca nbe recommendation or assistance | CDSS Purpose-Domain<br>Ontology Purpose |
| Diarrhea management | | |
| Disease management | | |
| DO | | Ontology Sources |
| domain | | |
| Domain experts | | |
| Drools | Workflow engine-Drools-flow | Rule Engine |
| DrugBank | | Ontology Sources |
| drug-drug interaction | | |
| Dublin core | | |
| dyslipidemia | | |
| EHIR rule engine | | |
| Emergency department | | |
| Emergency room | Or emergencey department | CDSS Purpose-Domain |
| Euler/EYE inference engine | | |
| evaluation | | |
| Expert | | CDSS Rule Sources<br>Ontology Sources |
| expert system | | |
| FaCTþþsemantic reasoner | | |
| Fall prevention | | |

| | | |
|---|---|---|
| Feasibility | | CDSS Evaluation (who, what, how, where) |
| Feasibility study | | |
| FHIR | | |
| Follow up care | | CDSS Purpose-Domain |
| Foundational Model of Anatomy (FMA | | |
| Framework | | |
| Framework\| | | |
| FSM(Finite State Machine) based execution engine | | |
| Fuzzy logic inference | | |
| Fuzzy rules | | |
| Galen Ontology | | Ontology Sources |
| Gastric cancer | | |
| Gastroenterologist | | |
| GELLO | | |
| GEM execution engine | | Rule Engine |
| GEM: the Guideline Elements Model | | CDSS Rule Sources |
| General practitioner | | |
| Guidelines | | CDSS Rule Sources<br>Ontology Sources |
| Guidelines in GLIF3 |  | CDSS Rule Sources |
| Head trauma | | |
| Heart failure | | CDSS Purpose-Domain |
| Hemodialysis | | |
| HL7 | | |
| HPO | | Ontology Sources |
| Hypertension | | CDSS Purpose-Domain |
| ICD-10 | | Ontology Sources |
| ICD-9-CM | | |
| ICU | | CDSS Purpose-Domain |
| IDO | | Ontology Sources |
| ILOG JRules | | |
| Images | | |
| implementation | | |
| Infectious disease | | CDSS Purpose-Domain |
| Inference engine | | Rule Engine |
| inference rule | | |

| | | |
|---|---|---|
| Inference/reasoning | It can include recommendations, to get implicit relationships, assessment, Infer context information, patient conditions; Infer clinical tests on patients to detect AD | CDSS Rule Sources<br>Ontology Purpose |
| inference-based | | |
| inpatient | | CDSS Purpose-Domain |
| JADE | | |
| Java | | |
| JBoss rule | | |
| Jena | | |
| JENA inference engine | Jena reasoner | Rule Engine |
| Jena Reasoner | | |
| JENA rule engine | | Rule Engine |
| JENA rules | | CDSS Rule Format |
| Jess reasoner | | Rule Engine |
| Jess Rule Engine | | Rule Engine |
| Jess rules | | CDSS Rule Format |
| JRules | including Jrules OWL plug-in | Rule Management<br>CDSS Rule Author/Edit |
| JRules OWL plug-in | | |
| JSON | | |
| Knowledge source | It can incldue CDSS rule source; Derive data, support, interface, configuration models | Ontology Purpose |
| Lab | | |
| language | | |
| Lipid management | | CDSS Purpose-Domain |
| Literature | | CDSS Rule Sources<br>Ontology Sources |
| logic element (class) | | |
| LOINC | | Ontology Sources |
| lung cancer | | |
| Medical records | Also can be patient records, patient cases, can be identified or de-identified | CDSS Rule Sources<br>Ontology Sources<br>CDSS Evaluation (who, what, how, where) |
| medication order | | |

| | | |
|---|---|---|
| MLM | | |
| Natural Rule Language (NRL) | | |
| NCBI taxonomy | | Ontology Sources |
| NCI Terminology | | Ontology Sources |
| Nehprology | | |
| Neonatal | | |
| Neurosurgery | | |
| NICE guideline | | CDSS Rule Sources |
| notification | | |
| Object Contraint Language (OCL) | | |
| Object Management Group (OMG) | | |
| Oncology | | |
| ontology | | |
| Ontology inference performance | | CDSS Evaluation (who, what, how, where) |
| Ontology is validated by domain experts | | Ontology Validation By |
| OPCS | | |
| operation rule | | |
| Operational risk managment | | |
| Outpatient | | CDSS Purpose-Domain |
| OWL | | Ontology Language<br>CDSS Rule Format |
| patient | | |
| patient data | | |
| patient input | | |
| patient management | | |
| patient monitoring | | |
| patient safety | | |
| Patients' records | This is for CDSS Evaluation that conducted via using patient's records retrospectively. | CDSS Rule Sources<br>CDSS Evaluation (who, what, how, where) |
| Pattern matching | ? | Rule Engine |
| Pediatric | | |
| Pediatric orthopaedist | | |
| Pellet reasoner | | |
| pneumonia | | |
| Polypharmacy prescription | | CDSS Purpose-Domain<br>Ontology Purpose<br>CDSS Purpose-Type |
| Postoperative managment | | CDSS Purpose-Domain |
| Postoperative reports | | |

| | | |
|---|---|---|
| Prediction models/rules | Alvarado Score can be an example; also should include prediction models | CDSS Purpose-Domain<br>CDSS Purpose-Type |
| Preoperative | | |
| Preoperative assessment | | |
| Preoperative risk assessment | | CDSS Purpose-Domain |
| Prescription | | |
| Primary care providers | PCP can be a user group; can be knowledge source; can be an evaluator; can validate clinical knowledge. | CDSS Purpose-Domain |
| Production system | Including EMR, HIS, or EHR | Architeture Design |
| PROforma-based authoring | | |
| Prognosis | | |
| Prostatectomy | | |
| Protege | It can be used as a visual editing tool for OWL rules; including Protégé SWRL tab, Protégé + SWRLJessTab, | Rule Management<br>CDSS Rule Author/Edit |
| Prototype | | Architeture Design |
| provider input | | |
| Publication year | This is also used to record the paper. | |
| purpose | | |
| RacerPro reasoner | | |
| RDF/JSON | | |
| Reasoner: COSI (Closed World OWL Interpreter) | | Rule Engine |
| Recommendations | It can be recommendations for care managment, diagnosis, treatment or progrnosis | Ontology Purpose<br>CDSS Purpose-Type |

| | | |
|---|---|---|
| Reminder | It can be screening reminder, immunization, or other preventive service reminder | Ontology Purpose<br>CDSS Purpose-Type |
| Risk assessment | | |
| risk management | | |
| Rule authoring interface | | Rule Management |
| rule engine JSR-94 | | |
| Rule-based reasoner | (e.g., Pellet) | Rule Engine |
| RxNorm | | |
| SAGE | | |
| Sage format | | |
| SAGE model | | |
| SEBASTIAN rules | | |
| Self-managment | | |
| sepsis | | |
| Services on clinical data repository | This is a code for architecture mechanism for CDSS | Architeture Design |
| Simulated clinical setting for testing | | |
| SNOMED CT | | Ontology Sources |
| SPARQL | | |
| Speech language therapy | | |
| statistics-based | | |
| Suggestion | | |
| Surgery | | |
| SWAN | | Ontology Sources |
| SWRL rules | | CDSS Rule Format |
| Telemedicine system | | CDSS Purpose-Domain |
| temporal model | | |
| Test results-lab | | |
| tool | | |
| Training system | | |
| Treatment | It can be a recommendation or assistance | CDSS Purpose-Domain<br>Ontology Purpose |
| triage | | |
| uEngine | | |
| UMLS | | Ontology Sources |
| Urology | | |
| Use cases | Can be ICU or other units; also called scenarios. | CDSS Evaluation (who, what, how, where) |
| validation | | |
| verification | | |

| | | |
|---|---|---|
| Warning | | |
| WDSL | | |
| Web resources | | CDSS Rule Sources<br>Ontology Sources |
| Web services | Including Web API | Architeture Design |
| xml | | CDSS Rule Format |

**Code Groups**

Architecture Design (diagram, description)
Architeture Design
CDSS Evaluation (who, what, how, where)
CDSS Purpose
CDSS Purpose-Domain
CDSS Purpose-Perioperative
CDSS Purpose-Type
CDSS Purpose-User
CDSS Rule
CDSS Rule Author/Edit
CDSS Rule Engine
CDSS Rule Format
CDSS Rule Management
CDSS Rule Sources
CDSS Rule Sources/Ontology Sources

CDSS Setting
Guideline Engine
Integrated Development Environment (IDE)
Language
Metadata
Ontology Evaluation
Ontology Language
Ontology Purpose
Ontology Sources
Ontology Validation By
Program Language
Reasoner
Rule Engine
Rule Management
Standards
