## Appendix 3 for "A systematic review of ontology-based clinical decision support system rules: usage, management, and interoperability"

**Appendix 3: List of selected papers included in this systematic literature review (*n* = 81)**

| Publishing year | Author | Paper title | Journal/source | PMID |
|---|---|---|---|---|
| 2000 | A Manjarrés Riesco, R Martínez Tomás, J Mira Mira | A customisable framework for the assessment of therapies in the solution of therapy decision tasks | Artif Intell Med. 2000 Jan;18(1):57-82 | 10606794 |
| 2000 | S S Abidi, S Manickam | Transforming XML-based electronic patient records for use in medical case based reasoning systems | Stud Health Technol Inform. 2000;77:709-13 | 11187645 |
| 2000 | De Clercq PA, Blom JA, Hasman A, Korsten HH | GASTON: an architecture for the acquisition and execution of clinical guideline-application tasks | Med Inform Internet Med. 2000 Oct-Dec;25(4):247-63 | 11198187 |
| 2000 | Payne TH, Savarino J, Marshall R, Hoey CT | Use of a clinical event monitor to prevent and detect medication errors | Proc AMIA Symp. 2000:640-4 | 11079962 |
| 2001 | Achour SL, Dojat M, Rieux C, Bierling P, Lepage E | A UMLS-based knowledge acquisition tool for rule-based clinical decision support system development | J Am Med Inform Assoc. 2001 Jul-Aug;8(4):351-60 | 11418542 |
| 2001 | Séroussi B, Bouaud J, Dréau H, Falcoff H, Riou C, Joubert M, Simon C, Simon G, Venot A | ASTI: a guideline-based drug-ordering system for primary care | Stud Health Technol Inform. 2001;84(Pt 1):528-32 | 11604796 |
| 2002 | Karadimas HC, Chailloleau C, Hemery F, Simonnet J, Lepage E | Arden/J: an architecture for MLM execution on the Java platform | J Am Med Inform Assoc. 2002 Jul-Aug;9(4):359-68 | 12087117 |
| 2002 | Ray HN, Boxwala AA, Anantraman V, Ohno-Machado L | Providing context-sensitive decision-support based on WHO guidelines | Proc AMIA Symp. 2002:637-41 | 12463901 |
| 2002 | Das AK, Musen MA | SYNCHRONUS: a reusable software module for temporal integration | Proc AMIA Symp. 2002:195-9 | 12463814 |
| 2003 | Poon EG, Wang SJ, Gandhi TK, Bates DW, Kuperman GJ | Design and implementation of a comprehensive outpatient Results Manager | J Biomed Inform. 2003 Feb-Apr;36(1-2):80-91 | 14552849 |
| 2003 | Siaw-Teng Liaw, Nabil Sulaiman, Christopher Pearce, Jane Sims, Keith Hill, Heather Grain, Justin Tse, Choon-Kiat Ng | Falls prevention within the Australian general practice data model: methodology, information model, and terminology issues | J Am Med Inform Assoc. Sep-Oct 2003;10(5):425-32 | 12807809 |

| | | | | |
|---|---|---|---|---|
| 2003 | Barth C, Tobman M, Nätscher C, Sussmann H, Horsch A | Fusing a systematic and a case-based repository for medical decision support | Stud Health Technol Inform. 2003;95:560-4 | 14664046 |
| 2004 | Greenes RA, Sordo M, Zaccagnini D, Meyer M, Kuperman GJ | Design of a standards-based external rules engine for decision support in a variety of application contexts: report of a feasibility study at Partners HealthCare System | Stud Health Technol Inform. 2004;107(Pt 1):611-5 | 15360885 |
| 2006 | Ebrahiminia V, Riou C, Seroussi B, Bouaud J, Dubois S, Falcoff H, Venot A | Design of a decision support system for chronic diseases coupling generic therapeutic algorithms with guideline-based specific rules | Stud Health Technol Inform. 2006;124:483-8. | 17108565 |
| 2006 | Kashyap V, Morales A, Hongsermeier T | On implementing clinical decision support: achieving scalability and maintainability by combining business rules and ontologies | AMIA Annu Symp Proc. 2006;2006:414-8 | 17238374 |
| 2007 | K. Verlaene, W. Joosen and P. Verbaeten | Arriclides: An Architecture Integrating Clinical Decision Support Models | HICSS '07: Proceedings of the 40th Annual Hawaii International Conference on System Sciences January 2007, pp 135 | ACM |
| 2007 | S. R. Abidi | Ontology-Based Modeling of Breast Cancer Follow-up Clinical Practice Guideline for Providing Clinical Decision Support | CBMS '07: Proceedings of the Twentieth IEEE International Symposium on Computer-Based Medical Systems June 2007, pp 542–547 | ACM |
| 2007 | P Jannin, X Morandi | Surgical models for computer-assisted neurosurgery | Neuroimage. 2007 Sep 1;37(3):783-91 | 17613249 |
| 2007 | Michael Stacey, Carolyn McGregor, Mark Tracy | An architecture for multi-dimensional temporal abstraction and its application to support neonatal intensive care | Annu Int Conf IEEE Eng Med Biol Soc. 2007;2007:3752-6 | 18002814 |
| 2007 | Papageorgiou E, Stylios C, Groumpos P | Novel Architecture for supporting medical decision making of different data types based on Fuzzy Cognitive Map Framework | Annu Int Conf IEEE Eng Med Biol Soc. 2007;2007:1192-5 | 18002176 |
| 2008 | Chiara Cornalba, Roberto G Bellazzi, Riccardo Bellazzi | Building a normative decision support system for clinical and operational risk management in hemodialysis | IEEE Trans Inf Technol Biomed. 2008 Sep;12(5):678-86 | 18779083 |
| 2009 | Farion K, Michalowski W, Wilk S, O'Sullivan D, Rubin S, Weiss D | Clinical decision support system for point of care use--ontology-driven design and software implementation | Methods Inf Med. 2009;48(4):381-90 | 19448882 |

| | | | | |
|---|---|---|---|---|
| 2009 | Tien Tuan Dao, Frédéric Marin, Marie Christine Ho Ba Tho | Clinical validated Computer-Aided Decision System to the clubfeet deformities | Annu Int Conf IEEE Eng Med Biol Soc. 2009;2009:6230-3 | 19965086 |
| 2009 | Q. Zhou | A Clinical Decision Support System for Metabolism Synthesis | CINC '09: Proceedings of the 2009 International Conference on Computational Intelligence and Natural Computing - Volume 02 June 2009, pp 323–325 | ACM |
| 2009 | Carenini M; ReMINE Consortium | ReMINE: an ontology-based risk management platform | Stud Health Technol Inform. 2009;148:32-42 | 19745233 |
| 2010 | Lee J, Kim J, Cho I, Kim Y | Integration of workflow and rule engines for clinical decision support services | Stud Health Technol Inform. 2010;160(Pt 2):811-5 | 20841798 |
| 2010 | F. Ongenae, T. Dhaene, F. D. Turck, D. Benoit and J. Decruyenaere | Design of a probabilistic ontology-based clinical decision support system for classifying temporal patterns in the ICU: A sepsis case study | CBMS '10: Proceedings of the 2010 IEEE 23rd International Symposium on Computer-Based Medical Systems October 2010, pp 389–394 | ACM |
| 2010 | Basilakis J, Lovell NH, Redmond SJ, Celler BG | Design of a decision-support architecture for management of remotely monitored patients | IEEE Trans Inf Technol Biomed. 2010 Sep;14(5):1216-26 | 20615815 |
| 2010 | Szymon Wilk, Wojtek Michalowski, Ken Farion, Jelber Sayyad Shirabad | MET3-AE system to support management of pediatric asthma exacerbation in the emergency department | Stud Health Technol Inform. 2010;160(Pt 2):841-5 | 20841804 |
| 2010 | Borbolla D, Otero C, Lobach DF, Kawamoto K, Gomez Saldaño AM, Staccia G, Lopez G, Figar S, Luna D, Bernaldo de Quiros FG | Implementation of a clinical decision support system using a service model: results of a feasibility study | Stud Health Technol Inform. 2010;160(Pt 2):816-20 | 20841799 |
| 2011 | M.-M. Bouamrane, A. Rector and M. Hurrell | Using OWL ontologies for adaptive patient information modelling and preoperative clinical decision support | Knowledge and Information Systems (KAIS), Volume 29, Issue 2 November 2011, pp 405–418 | ACM |

| | | | | |
|---|---|---|---|---|
| 2011 | Tien Tuan Dao, Frédéric Marin, Henri Bensahel, Marie Christine Ho Ba Tho | Computer-aided decision system for the clubfeet deformities | Adv Exp Med Biol. 2011;696:623-35 | 21431604 |
| 2011 | Cao F, Sun X, Wang X, Li B, Li J, Pan Y | Ontology-based knowledge management for personalized adverse drug events detection | Stud Health Technol Inform. 2011;169:699-703 | 21893837 |
| 2011 | Chang-Shing Lee, Mei-Hui Wang | A fuzzy expert system for diabetes decision support application | IEEE Trans Syst Man Cybern B Cybern. 2011 Feb;41(1):139-53 | 20501347 |
| 2012 | Riaño D, Real F, López-Vallverdú JA, Campana F, Ercolani S, Mecocci P, Annicchiarico R, Caltagirone C | An ontology-based personalization of health-care knowledge to support clinical decisions for chronically ill patients | J Biomed Inform. 2012 Jun;45(3):429-46 | 22269224 |
| 2012 | Bright TJ, Yoko Furuya E, Kuperman GJ, Cimino JJ, Bakken S | Development and evaluation of an ontology for guiding appropriate antibiotic prescribing | J Biomed Inform. 2012 Feb;45(1):120-8 | 22019377 |
| 2012 | Koutkias V, Kilintzis V, Stalidis G, Lazou K, Niès J, Durand-Texte L, McNair P, Beuscart R, Maglaveras N | Knowledge engineering for adverse drug event prevention: on the design and development of a uniform, contextualized and sustainable knowledge-based framework | J Biomed Inform. 2012 Jun;45(3):495-506 | 22326287 |
| 2012 | Grando A, Farrish S, Boyd C, Boxwala A | Ontological approach for safe and effective polypharmacy prescription | AMIA Annu Symp Proc. 2012;2012:291-300. Epub 2012 Nov 3 | 23304299 |
| 2012 | Chniti A, Boussadi A, Degoulet P, Albert P, Charlet J | Pharmaceutical validation of medication orders using an OWL Ontology and Business Rules | Stud Health Technol Inform. 2012;180:1224-6 | 22874408 |
| 2012 | Paterno MD, Goldberg HS, Simonaitis L, Dixon BE, Wright A, Rocha BH, Ramelson HZ, Middleton B | Using a service oriented architecture approach to clinical decision support: performance results from two CDS Consortium demonstrations | AMIA Annu Symp Proc. 2012;2012:690-8. Epub 2012 Nov 3 | 23304342 |
| 2013 | Haug PJ, Ferraro JP, Holmen J, Wu X, Mynam K, Ebert M, Dean N, Jones J | An ontology-driven, diagnostic modeling system | J Am Med Inform Assoc. 2013 Jun;20(e1):e102-10 | 23523876 |

| | | | | |
|---|---|---|---|---|
| 2013 | W. Yao and A. Kumar | CONFlexFlow: Integrating Flexible clinical pathways into clinical decision support systems using context and rules | Decision Support Systems (DSSY), Volume 55, Issue 2 May, 2013, pp 499–515 | ACM |
| 2013 | A. Artetxe, E. Sanchez, C. Toro, C. Sanín, E. Szczerbicki, M. Graña and J. Posada | Impact of reflexive ontologies in semantic clinical decision support systems | Cybernetics and Systems (CSYS), Volume 44, Issue 2-3 March 2013, pp 187–203 | ACM |
| 2013 | Farkash A, Timm JT, Waks Z | A model-driven approach to clinical practice guidelines representation and evaluation using standards | Stud Health Technol Inform. 2013;192:200-4 | 23920544 |
| 2013 | Sáez C, Bresó A, Vicente J, Robles M, García-Gómez JM | An HL7-CDA wrapper for facilitating semantic interoperability to rule-based Clinical Decision Support Systems | Comput Methods Programs Biomed. 2013 Mar;109(3):239-49 | 23199936 |
| 2013 | Corrigan D, Taweel A, Fahey T, Arvanitis T, Delaney B | An ontological treatment of clinical prediction rules implementing the Alvarado score | Stud Health Technol Inform. 2013;186:103-7 | 23542977 |
| 2013 | Reza Shojanoori, Radmila Juric | Semantic remote patient monitoring system | Telemed J E Health. 2013 Feb;19(2):129-36 | 23363406 |
| 2013 | S Wilk, W Michalowski, D O'Sullivan, K Farion, J Sayyad-Shirabad, C Kuziemsky, B Kukawka | A task-based support architecture for developing point-of-care clinical decision support systems for the emergency department | Methods Inf Med. 2013;52(1):18-32 | 23232759 |
| 2013 | Özgün Yılmaz, Rıza Cenk Erdur, Mustafa Türksever | SAMS--a systems architecture for developing intelligent health information systems | J Med Syst. 2013 Dec;37(6):9989 | 24197356 |
| 2014 | Bau CT, Chen RC, Huang CY | Construction of a clinical decision support system for undergoing surgery based on domain ontology and rules reasoning | Telemed J E Health. 2014 May;20(5):460-72 | 24730353 |
| 2014 | Hua-qiong Wang, Tian-shu Zhou, Li-li Tian, Yang-ming Qian, Jing-song Li | Creating hospital-specific customized clinical pathways by applying semantic reasoning to clinical data | J Biomed Inform. 2014 Dec;52:354-63 | 25109270 |
| 2014 | Sesen MB, Peake MD, Banares-Alcantara R, Tse D, Kadir T, Stanley R, Gleeson F, Brady M | Lung Cancer Assistant: a hybrid clinical decision support application for lung cancer care | J R Soc Interface. 2014 Sep 6;11(98):20140534 | 24990290 |
| 2014 | M. Gallerani, D. Pelizzola, M. Pivanti, M. | Appropriateness of Repeated Execution of Laboratory Examinations: A CDSS Approach | ICHI '14: Proceedings of the 2014 IEEE International Conference on Healthcare | ACM |

| | | | | |
|---|---|---|---|---|
| | Boni, E. Lamma and E. Bellodi | | Informatics September 2014, pp 161–169 | |
| 2014 | Goldberg HS, Paterno MD, Rocha BH, Schaeffer M, Wright A, Erickson JL, Middleton B | A highly scalable, interoperable clinical decision support service | J Am Med Inform Assoc. 2014 Feb;21(e1):e55-62 | 23828174 |
| 2014 | Samuel Alan Stewart, Samina Abidi, Louise Parker, Mark Bernstein, Syed Sibte Raza Abidi | Clinical guideline-driven personalized self-management diary for paediatric cancer survivors | Stud Health Technol Inform. 2014;205:18-22 | 25160137 |
| 2015 | Ying Shen, Joël Colloc, Armelle Jacquet-Andrieu, Kai Lei | Emerging medical informatics with case-based reasoning for aiding clinical decision in multi-agent system | J Biomed Inform. 2015 Aug;56:307-17 | 26133480 |
| 2015 | Brendan C Delaney, Vasa Curcin, Anna Andreasson, Theodoros N Arvanitis, Hilde Bastiaens, Derek Corrigan, Jean-Francois Ethier, Olga Kostopoulou, Wolfgang Kuchinke, Mark McGilchrist, Paul van Royen, Peter Wagner | Translational Medicine and Patient Safety in Europe: TRANSFoRm--Architecture for the Learning Health System in Europe | Biomed Res Int. 2015;2015:961526 | 26539547 |
| 2015 | Borna Jafarpour, Samina Raza Abidi, Ahmad Marwan Ahmad, Syed Sibte Raza Abidi | INITIATE: An Intelligent Adaptive Alert Environment | Stud Health Technol Inform. 2015;216:285-9 | 26262056 |
| 2015 | Vladimir Robles-Bykbaev, Martín López-Nores, José Pazos-Arias, Diego Quisi-Peralta, Jorge García-Duque | An Ecosystem of Intelligent ICT Tools for Speech-Language Therapy Based on a Formal Knowledge Model | Stud Health Technol Inform. 2015;216:50-4 | 26262008 |
| 2016 | Szymon Wilk, Mounira Kezadri-Hamiaz, Daniela Rosu, Craig Kuziemsky, Wojtek Michalowski, | Using Semantic Components to Represent Dynamics of an Interdisciplinary Healthcare Team in a Multi-Agent Decision Support System | J Med Syst. 2016 Feb;40(2):42 | 26590980 |

| | | | | |
|---|---|---|---|---|
| | Daniel Amyot, Marc Carrier | | | |
| 2016 | Samina Raza Abidi, Jafna Cox, Ashraf Abusharekh, Nima Hashemian, Syed Sibte Raza Abidi | A Digital Health System to Assist Family Physicians to Safely Prescribe NOAC Medications | Stud Health Technol Inform. 2016;228:519-23 | 27577437 |
| 2016 | Zhang YF, Tian Y, Zhou TS, Araki K, Li JS | Integrating HL7 RIM and ontology for unified knowledge and data representation in clinical decision support systems | Comput Methods Programs Biomed. 2016 Jan;123:94-108 | 26474836 |
| 2016 | Goldberg HS, Paterno MD, Grundmeier RW, Rocha BH, Hoffman JM, Tham E, Swietlik M, Schaeffer MH, Pabbathi D, Deakyne SJ, Kuppermann N, Dayan PS | Use of a remote clinical decision support service for a multicenter trial to implement prediction rules for children with minor blunt head trauma | Int J Med Inform. 2016 Mar;87:101-10 | 26806717 |
| 2016 | Luis Marco-Ruiz, Carlos Pedrinaci, J A Maldonado, Luca Panziera, Rong Chen, J Gustav Bellika | Publication, discovery and interoperability of Clinical Decision Support Systems: A Linked Data approach | J Biomed Inform. 2016 Aug;62:243-64 | 27401856 |
| 2017 | Zhang YF, Gou L, Zhou TS, Lin DN, Zheng J, Li Y, Li JS | An ontology-based approach to patient follow-up assessment for continuous and personalized chronic disease management | J Biomed Inform. 2017 Aug;72:45-59 | 28676255 |
| 2017 | Shang Y, Wang Y, Gou L, Wu C, Zhou T, Li JS | Development of a Service-Oriented Sharable Clinical Decision Support System Based on Ontology for Chronic Disease | Stud Health Technol Inform. 2017;245:1153-1157 | 29295283 |
| 2017 | Chen RC, Jiang HQ, Huang CY, Bau CT | Clinical Decision Support System for Diabetes Based on Ontology Reasoning and TOPSIS Analysis | J Healthc Eng. 2017;2017:4307508 | 29312655 |
| 2017 | Kopanitsa G | Integration of Hospital Information and Clinical Decision Support Systems to Enable the Reuse of Electronic Health Record Data | Methods Inf Med. 2017 May 18;56(3):238-247 | 28361157 |
| 2017 | Samina Abidi | A Knowledge-Modeling Approach to Integrate Multiple Clinical Practice Guidelines to Provide Evidence-Based Clinical Decision Support for Managing Comorbid Conditions | J Med Syst. 2017 Oct 26;41(12):193 | 29076113 |
| 2018 | Shen Y, Yuan K, Chen D, Colloc J, Yang M, Li Y, Lei K | An ontology-driven clinical decision support system (IDDAP) for infectious disease diagnosis and antibiotic prescription | Artif Intell Med. 2018 Mar;86:20-32 | 29433958 |

| | | | | |
|---|---|---|---|---|
| 2018 | Hirenkumar Nakawala, Giancarlo Ferrigno, Elena De Momi | Development of an intelligent surgical training system for Thoracentesis | Artif Intell Med. 2018 Jan;84:50-63 | 29169646 |
| 2018 | Brigitte Séroussi, Gilles Guézennec, Jean-Baptiste Lamy, Naiara Muro, Nekane Larburu, Booma Devi Sekar, Coralie Prebet, Jacques Bouaud | Reconciliation of multiple guidelines for decision support: a case study on the multidisciplinary management of breast cancer within the DESIREE project | AMIA Annu Symp Proc. 2018 Apr 16;2017:1527-1536 | 29854222 |
| 2018 | Winter A, Stäubert S, Ammon D, Aiche S, Beyan O, Bischoff V, Daumke P, Decker S, Funkat G, Gewehr JE, de Greiff A, Haferkamp S, Hahn U, Henkel A, Kirsten T, Klöss T, Lippert J, Löbe M, Lowitsch V, Maassen O, Maschmann J, Meister S, Mikolajczyk R, Nüchter M, Pletz MW, Rahm E, Riedel M, Saleh K, Schuppert A, Smers S, Stollenwerk A, Uhlig S, Wendt T, Zenker S, Fleig W, Marx G, Scherag A, Löffler M | Smart Medical Information Technology for Healthcare (SMITH) | Methods Inf Med. 2018 Jul;57(S 01):e92-e105 | 30016815 |
| 2018 | Brigitte Seroussi, Jean-Baptiste Lamy, Naiara Muro, Nekane Larburu, Booma Devi Sekar, Gilles Guézennec, Jacques Bouaud | Implementing Guideline-Based, Experience-Based, and Case-Based Approaches to Enrich Decision Support for the Management of Breast Cancer Patients in the DESIREE Project | Stud Health Technol Inform. 2018;255:190-194 | 30306934 |
| 2018 | Wenquan Jin, Do Hyeun Kim | Design and Implementation of e-Health System Based on Semantic Sensor Network Using IETF YANG | Sensors (Basel). 2018 Feb 20;18(2):629 | 29461493 |

| | | | | |
|---|---|---|---|---|
| 2019 | Román-Villarán E, Pérez-Leon FP, Escobar-Rodriguez GA, Martínez-García A, Álvarez-Romero C, Parra-Calderón CL | An Ontology-Based Personalized Decision Support System for Use in the Complex Chronically Ill Patient | Stud Health Technol Inform. 2019 Aug 21;264:758-762 | 31438026 |
| 2019 | Semenov I, Osenev R, Gerasimov S, Kopanitsa G, Denisov D, Andreychuk Y | Experience in Developing an FHIR Medical Data Management Platform to Provide Clinical Decision Support | Int J Environ Res Public Health. 2019 Dec 20;17(1):73 | 31861851 |
| 2019 | Nguyen BP, Reese T, Decker S, Malone D, Boyce RD, Beyan O | Implementation of Clinical Decision Support Services to Detect Potential Drug-Drug Interaction Using Clinical Quality Language | Stud Health Technol Inform. 2019 Aug 21;264:724-728 | 31438019 |
| 2019 | Borna Jafarpour, Samina Raza Abidi, William Van Woensel, Syed Sibte Raza Abidi | Execution-time integration of clinical practice guidelines to provide decision support for comorbid conditions | Artif Intell Med. 2019 Mar;94:117-137 | 30871678 |
| 2019 | El-Sappagh S, Ali F, Hendawi A, Jang JH, Kwak KS | A mobile health monitoring-and-treatment system based on integration of the SSN sensor ontology and the HL7 FHIR standard | BMC Med Inform Decis Mak. 2019 May 10;19(1):97 | 31077222 |
| 2020 | Maldonado JA, Marcos M, Fernández-Breis JT, Giménez-Solano VM, Legaz-García MDC, Martínez-Salvador B. | CLIN-IK-LINKS: A platform for the design and execution of clinical data transformation and reasoning workflows. | Comput Methods Programs Biomed. 2020;**197**:105616 | 32629294 |
